# Evaluating the risk of mosquito-borne diseases in non-endemic regions: A dynamic modeling approach

**DOI:** 10.1101/2024.10.10.24315163

**Authors:** Nico Stollenwerk, Luís Mateus, Vanessa Steindorf, Bruno V. Guerrero, Rubén Blasco-Aguado, Aitor Cevidanes, Joseba Bidaurrazaga Van-Dierdonck, Maíra Aguiar

**Affiliations:** Basque Center for Applied Mathematics (BCAM), Bilbao, Spain; Center for Mathematical Studies (CEMS.UL) - Faculty of Sciences of the University of Lisbon (FCUL), Lisbon, Portugal; Scuola Superiore Meridionale (SSM), Naples, Italy; Animal Health Department, NEIKER - Basque Institute for Agricultural Research and Development, Basque Research and Technology Alliance (BRTA), Derio, Bizkaia, Spain; Public Health, Basque Health Department, Rekalde Zumarkalea 39A, 48008 Bilbao, Spain; Ikerbasque, Basque Foundation for Science, Bilbao, Spain

## Abstract

Mosquito-borne diseases are spreading into temperate zones, raising concerns about local outbreaks driven by imported cases. Using stochastic methods, we developed a vector-host model to estimate the risk of import-driven autochthonous outbreaks in non-endemic regions. The model explores key factors such as imported cases and vector abundance. Our analysis shows that mosquito population abundance significantly affects the probability and timing of outbreaks. Even with moderate mosquito populations, isolated or clustered outbreaks can be triggered, highlighting the importance of monitoring vector abundance for effective public health planning and interventions.

## 1 Introduction

The presence of *Aedes* mosquitoes in non-endemic regions has become a significant public health concern [1, 2], as these areas now face the potential risk of local transmission of mosquito-borne diseases such as dengue fever, Zika virus disease, and chikungunya fever [3]. Historically confined to tropical and subtropical climates, these diseases are now spreading into temperate zones, driven by factors like climate change, increased global travel, and the adaptability of these mosquitoes [4], which are now distributed across several parts of the world [5].

The expansion of mosquito species into new regions creates an alarming epidemiological situation, referred to as an “invasion scenario of disease introduction”. This scenario represents an increased risk of mosquito-borne disease cases transmitted by newly established mosquito species that serve as disease vectors, going beyond a small local outbreak or merely reaching a threshold for continuous transmission. Such a situation presents new challenges for public health systems in non-endemic areas, which may not be adequately equipped to manage these emerging disease outbreaks.

Understanding the concept of invasion scenarios of disease introduction is crucial for assessing the risk of autochthonous (locally transmitted) disease outbreaks in non-endemic regions [6, 7, 8, 9]. Even when a region is below the epidemiological threshold - indicating that the disease is not yet self-sustaining - significant outbreaks of autochthonous cases can still occur. Important examples include the Madeira Island outbreak in Portugal in 2012 [10] and more recent cases in France and Italy [11].

In these invasion scenarios, the risk of autochthonous cases is closely linked to both the abundance of mosquito vectors and the influx of viremic imported cases - individuals who are infected returning from travels in endemic areas who are capable of transmitting the virus [12, 13, 14, 15]. As mosquito populations increase, the likelihood of disease outbreaks also rises, often following power-law scaling in the distribution of confirmed cases [16, 17, 18]. This pattern, characteristic of systems approaching a critical threshold, indicates that small changes in key factors like mosquito population density or environmental conditions can lead to disproportionately large effects [19]. Recognizing this is essential for public health, emphasizing the need for early intervention, mosquito control, and continuous surveillance to prevent the system from reaching a tipping point where large-scale outbreaks become inevitable.

In the epidemiology of mosquito-borne diseases, classical compartmental models, both with and without explicit mosquito dynamics, have been extensively used, primarily focusing on disease transmission in endemic regions [20, 21, 22, 23, 24]. These models provide essential parameters for understanding disease dynamics. However, assessing the potential impact of mosquito abundance and imported cases on invasion scenarios - where no autochthonous cases have yet occurred - presents additional challenges, such as data limitations, dynamic variability, and the stochastic nature of disease spread.

To evaluate and quantify the risk of imported driven autochthonous cases in non-endemic regions, we propose a dynamic modeling approach that incorporates factors such as the presence of *Aedes* mosquitoes and viremic imported cases. Using stochastic methods and their deterministic counterpart, this paper offers a comprehensive analysis of the risk landscape, including estimates of waiting times until the first autochthonous case is identified and statistics on the inter-event times of imported cases, with confidence intervals derived from likelihood functions. Using the Basque Country as a case study - where mosquito populations are established and imported cases are frequently reported - we analyze trends in imported cases and assess the impact of mosquito abundance on the risk of autochthonous disease outbreaks. Our study provides a timely framework for predicting imported driven autochthonous disease cases based on available data on mosquito presence and imported case notifications, which is crucial for preventing sustained transmission and mitigating the effects of isolated outbreaks in non-endemic regions.

## 2 The SIRUV modeling framework

To assess the risk of mosquito-borne disease transmission in non-endemic regions, we refined and extended the SIRUV modeling framework [25, 26]. Initially designed as a foundational model for simulating disease spread in endemic areas, the framework is adjusted to describe local transmission dynamics in non-endemic regions, potentially triggered by the introduction of viremic imported cases and the presence of competent mosquito vectors.

In this section, we start by outlining the structure and key assumptions of the deterministic SIRUV model, which captures the interaction between human and mosquito populations in an endemic scenario. Using parameters obtained from data from endemic countries [27, 28, 29], the model is then extended to incorporate imported cases - which have little impact on sustained transmission in endemic areas - lower vector abundance, and stochastic elements. This extension allows for a more comprehensive analysis of how these factors influence the probability of autochthonous disease outbreaks in non-endemic regions.

### 2.1 The SIRUV model for endemic areas

The classical Susceptible-Infected-Recovered (SIR) model [30, 24], which describes the dynamics of a host-host transmission disease, is given by

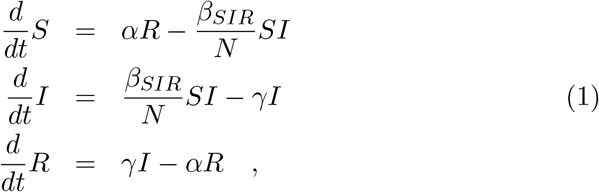

where *β*_*SIR*_ represents the transmission rate, *γ* the recovery rate, *N* the human population size and *α* the waning immunity rate. These equations describe the flow of individuals between the compartments: *S* (susceptible), *I* (infected), and *R* (recovered).

To model mosquito-borne diseases, the SIR model is extended to include explicit vector dynamics [31, 32, 25, 26, 33, 34, 35]. In its basic concept to describe different mosquito-borne diseases such as dengue, Zika and chikungunya, this results in the SIRUV type model, which incorporates compartments for Uninfected mosquitoes *U*, and infected mosquitoes, i.e., the disease-Vector *V*. The equations for the SIRUV model are given by

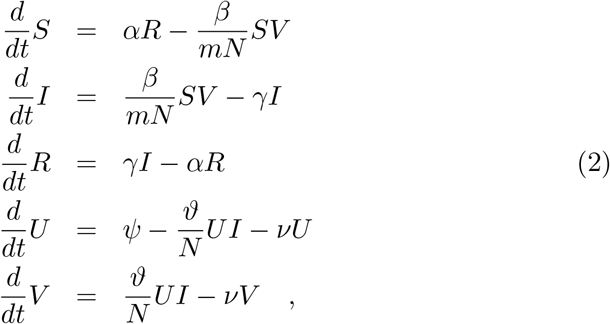

where *ψ* = *ν* · *mN* represents the mosquito supply rate, with *m* being the ratio of the mosquito population size to the human population size *N*, *ν* the natural mosquito mortality rate, and *β* and ϑ the transmission rates from mosquitoes to humans and humans to mosquitoes, respectively.

It is important to note that the average lifespan of an *Aedes* mosquito in nature is two weeks [36], whereas the protection conferred by human immunological responses persists for a much longer period. In the case of dengue fever, for example, protection against reinfection can last from 6 months to 2 years [28, 37, 38, 39, 40]. Given these differing timescales, a simplified model can be derived by using techniques such as singular perturbation or center manifold analysis [29, 26, 25] to describe time scale separation. As described in those studies, the mosquito dynamics are dominated by the slower dynamics of human infection on immunological time scales.

To rescale the mosquito parameters *ν* and ϑ to align with the human immunological timescales, we introduce a parameter *ϵ*, which represents the ratio between the fast mosquito timescale and the slow human timescale.

This rescaling is done such that 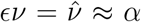 and 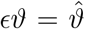. Hence, the last equation of system (2) becomes

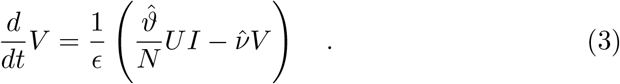

In the limit as *ϵ* → 0, indicating that mosquito dynamics quickly reach a quasi-stationary state, the equation simplifies to

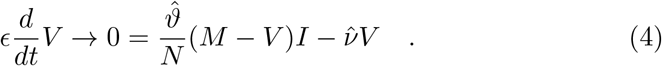

When considering the quasi-stationary solution for *V* (*I*) (i.e., 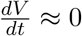), the number of infected mosquitoes *V* can be expressed as

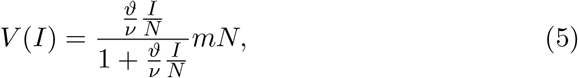

where *M* (*t*) := *U* (*t*) + *V* (*t*) represents the total mosquito population, which quickly approaches *M*^*^ = *mN*. This result captures how the number of infected mosquitoes adjusts according to the infection dynamics within the human population.

When the infected population is much smaller compared to the total population size *N*, as is the case for typical SIR systems in endemic areas, we have,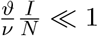 which simplifies equation (5) to

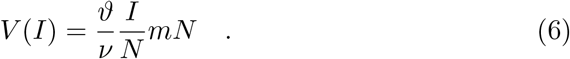

For more detailed calculations, see [29].

This simplified expression can be substituted for *V* in the human infection dynamics equations of system (2). Consequently, we obtain an effective SIR system with human-to-human transmission mediated by mosquitoes, given by

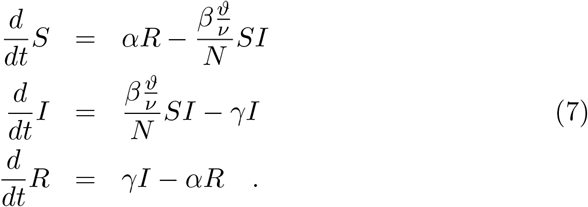

This relationship between models allows us to infer the basic infection parameters for mosquito-borne diseases from the host-host dynamic model given in system (1). Consequently, the infection rates from the SIRUV model and the effective SIR model are related by

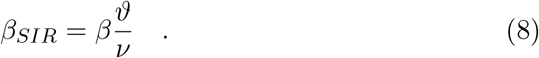

The parameter values used for our numerical simulations are listed in Table 1. Considering both primary and secondary infections, such as those observed in dengue, we set an approximate period of 10 years for the waning immunity *α*. The effective infectivity rate *β*_*SIR*_ is estimated from results in endemic regions, known to range from approximately 1.1 · *γ* in low endemic areas to 2 · *γ* in high endemic areas [27]. The selected value in our simulations reflects an effective infectivity rate, and it should be adjusted according to specific epidemiological information for the mosquito-borne disease and region under consideration. Any change in these values will be mentioned in the figure captions.

**Table 1:** Baseline model parameters to describe mosquito-borne disease transmission dynamics in endemic regions [27, 26]. These parameters can be refined according to specific epidemiological data from endemic areas and diseases.

| Parameter | Description | Value ($y^{-1}$) |
| --- | --- | --- |
| $\gamma$ | Recovery rate (1 week) | 52 |
| $\alpha$ | Waning immunity rate (10 years) | 0.1 |
| $\beta$ | Transmission rate from mosquitoes to humans | $1.2 \gamma$ |
| $\vartheta$ | Transmission rate from humans to mosquitoes | $1.2 \nu$ |
| $\nu$ | Mosquito mortality rate (10 days) | 36.5 |
| $\beta_{SIR}$ | Effective transmission rate | $1.2^2 \gamma$ |

### 2.2 The SIRUV model for an invasion scenario

To model an invasion scenario of disease introduction, we consider an area where there is no sustained local transmission, but where mosquitoes are present - which are significantly less abundant than in endemic areas - and imported cases are frequently recorded.

To quantify this relative reduction in the mosquito abundance, we introduce the ratio *k* ∈ [0, 1), defined as

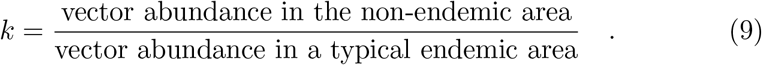

Since monitoring adult mosquitoes and effectively measuring their contribution to the infection process is challenging, the relative mosquito abundance *k*, could be measured using surrogate data such as ovitrap egg counts, positive ovitrap index, vector indices, composite index, passive larval and pupal collections, adult collections, and Stegomyia indices, for example [41, 42, 43].

The reduced supply rate of vectors *ψ*, in a non-endemic area can be expressed as a function of *k*

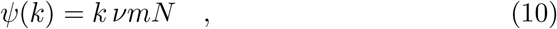

where *k* = 1 represents the vector supply rate typical of an endemic scenario.

On the other hand, the possibility of having imported infections *Y* into the population can be introduced in the model by assuming that there exists a steady input of new infections at constant ratio ϱ [28, 44]. In real life, those infections account for travelers arriving during their viremic period or residents returning from endemic regions with an active infection, which are often recorded in non-endemic areas when those individuals seek medical care. The number of imported cases is described by *Y* := ϱ*N*, with ϱ *>* 0, and it is included in the infection term of the uninfected mosquitoes equation.

After including these considerations in the System (2), the model for the non-endemic area reads

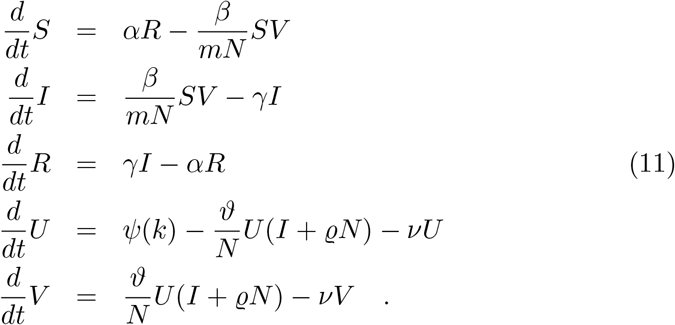

In this deterministic model, the expected number of imported cases is constant at ϱ*N*, which is relatively small compared to the total human population size (usually on the order of a few individuals). However, because infectious disease transmission is inherently stochastic, this variability must be considered in the modeling process.

### 2.3 The stochastic SIRUV model for an invasion scenario

The stochastic version of the SIRUV model given in system (11) is described by the dynamics of the discrete state vector <u>*X*</u> := (*S, I, R, U, V*)^tr^, representing the evolution of the state variables of the model. The probability of arriving to the state <u>*X*</u> at time *t* is modeled as a continuous time Markov process with the following master equation

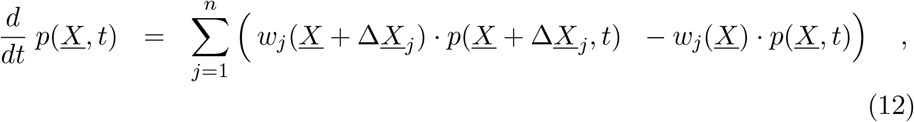

where *n* = 7 is the number of different deviations from state <u>*X*</u>, which are given by Δ<u>*X*</u>_*j*_ := *r*_*j*_. For the SIRUV model for the non-endemic scenario, the transitions *w*_*j*_(<u>*X*</u>) and their shift vectors <u>*r*</u>_*j*_ are

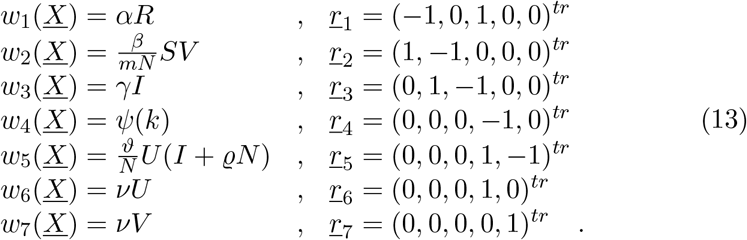

With these *w*_*j*_(*X*) and *r*_*j*_ specified, we can obtain stochastic realizations of the process using the Gillespie algorithm [45].

In the case of a well-defined system size, we can express the stochastic process in terms of the densities of the state variables. From this, we can obtain the mean field ODE system and, using the Kramers-Moyal approximation of the master equation, derive a Fokker-Planck equation and from it a system of stochastic differential equations [46]. In this study, we investigate stochastic processes with small numbers of cases, which are close to extinction thresholds. Therefore, we will primarily use the version with absolute numbers, rather than the densities.

The expected number of autochthonous infected cases is denoted by ⟨*I*⟩and is defined as

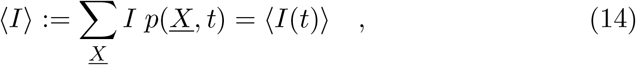

where *p*(*<u>X</u>, t*) represents the probability of the system being in state *X* at time *t*. The dynamics of ⟨*I*⟩ is governed by the evolution of these probabilities over time.

After performing the calculations, we obtain the following differential equation for the expected number of autochthonous infected cases

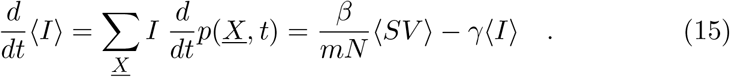

In the mean field approximation, this simplifies to

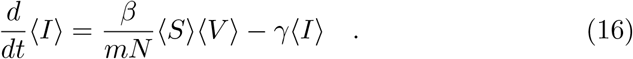

Thus, the ordinary differential equation (ODE) system captures the dynamics of the stochastic process in terms of expectation values [19].

### 2.4 Analytic results for the expected risk of autochthonous cases

After defining the SIRUV model for non-endemic areas and accounting for the stochastic nature of the process, we found that the expected number of autochthonous infected cases ⟨*I*⟩, depends on the number of imported cases *Y* = ϱ*N*, and the relative mosquito abundance *k*. We use the mean value of the quasi-stationary solution for *I* as a proxy to estimate the risk of autochthonous cases of arbovirus diseases in non-endemic regions.

From the invasion scenario model (see system (11)), which serves as a mean field approximation of the stochastic process, we derive an analytical expression for ⟨*I*⟩. For simplicity in notation, we will omit the expectation brackets in the following discussion.

Starting from the mosquito dynamics given by the last two equations in system (11), the dynamics of the total mosquito population *M* = *U* + *V*, is

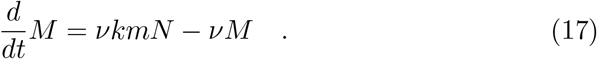

At stationarity, the total mosquito population is *M*^*^ = *kmN*, and the uninfected mosquito population is *U* = *M*^*^ − *V* = *kmN* − *V*.

Assuming the time scale separation 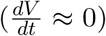 described in Section 2.1, the following expression for *V* (*I*) can be derived from the last equation in system (11)

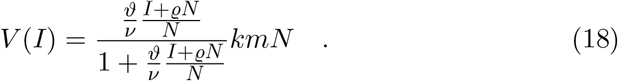

The assumption that 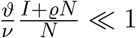 remains valid since the number of infected and imported cases is small compared to the total population. Consequently, the relationship between infected mosquitoes and infected humans is approximately linear and given by

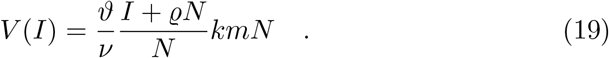

This expression can be substituted for *V* in the human disease dynamics equation in system (11)

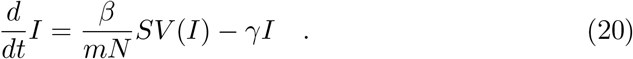

Considering that in a non-endemic area almost the entire population is susceptible, hence *S*(*t*) ≈ *S*_0_ ≈ *N*, the equation can be written as

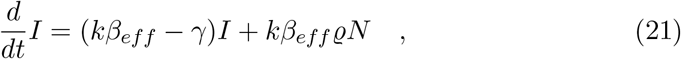

where 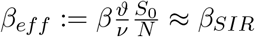 is the effective infection rate. Note that in the invasion scenario *k* can still modulate the infectivity.

For *kβ*_*eff*_ *< γ*, i.e., below the epidemiological threshold for exponential growth upon index cases (*β*_*eff*_ = *γ*), we have, triggered by import, a sub-critical stationary state of the dynamics given by Equation (21)

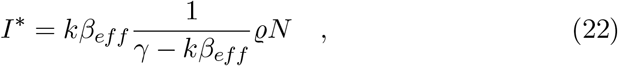

where *γ* − *kβ*_*eff*_ =: *ε*(*k*) is defined as the distance away from the exponential growth threshold with self-sustained spreading in the local population, depending on the relative mosquito abundance *k*. Hence, the expected number of autochthonous cases, given the underlying stochastic process is

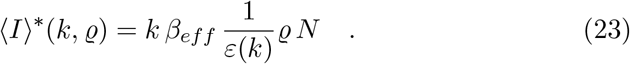

This expression depends on the mosquito abundance *k*, in the invasion region and the import of infection ϱ. Although ⟨*I*⟩ is typically much smaller than one case, stochastic simulations can reveal isolated autochthonous cases and occasionally rare clusters of outbreaks, where more than one case occurs simultaneously, as illustrated in Figure 1.

**Figure 1:**
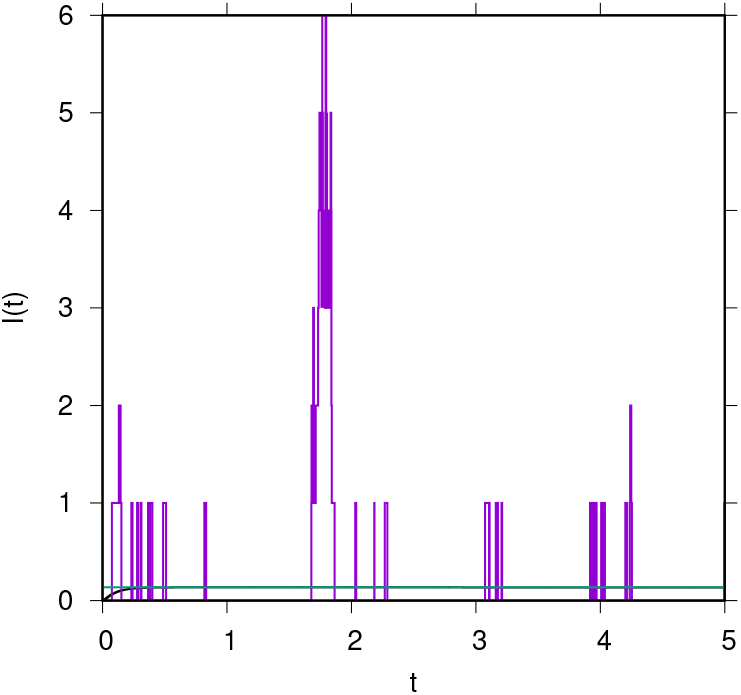
One stochastic realization (in magenta), the trajectory towards the stationary state of the SIRUV model (in black), and the approximation solution using the equation (in green) with *k* = 40% as the proportion of mosquitoes relative to endemic countries. For *N* = 10^4^ and ϱ = 10^−5^, the parameter values used in this simulation are listed in Table 1.

### 2.5 The SIRUV model with explicit dynamics for imported cases *Y*

Aimed at understanding the impact of imported cases on the dynamics of stochastic realizations, we extend the SIRUV framework by explicitly incorporating the dynamics of imported cases *Y*, to study their stochastic fluctuations.

The SIRUVY model can be expressed as

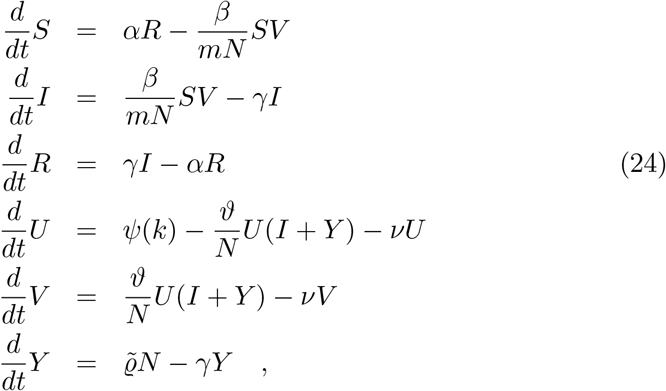

where 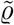 represents the constant input rate of imported cases, which can recover at rate *γ*. Note that the last equation is decoupled from System (24), and consequently, the stationary value of the imported cases *Y* ^*^, is given by

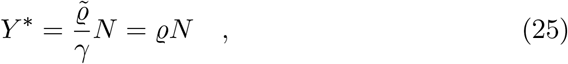

consistent with the previous model in System (11).

When considering the cumulative imported cases *C*_*Y*_, over a given time interval, we have

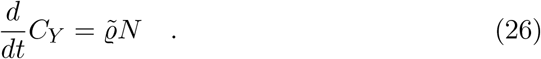

The numerical solutions for both the number of imported cases *Y* (*t*), and the cumulative imported cases *C*_*Y*_ (*t*), are illustrated in Figure 2. With initial conditions set to zero imported cases *Y* (*t*_0_) = 0, we observe that the mean field solution of the system described by Equation (24) converges quickly to its stationary state, as shown in Figure 2(a). After five years of simulation, the cumulative number of imported cases is expected to be approximately 26 cases, see Figure 2(b).

**Figure 2:**
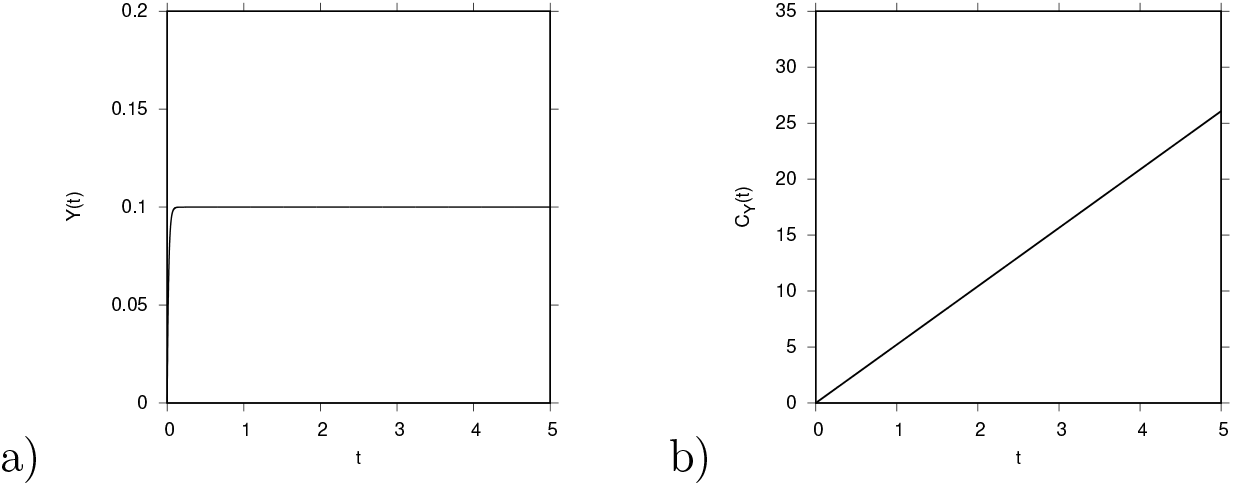
Expected values from the mean field solution over a 5-year simulation period. For *N* = 10^4^ and ϱ = 10^−5^, the parameter values used in this simulation are listed in Table 1. In (a), the imported cases and in (b), the cumulative cases *C*_*Y*_ (*t*).

### 2.6 The stochastic SIRUVY model

Following the stochastic approach described in Section 2.3, we model the SIRUVY system using the master equation (see equation (12)). The stochastic model is characterized by the extended state vector <u>*X*</u> := (*S, I, R, U, V, Y*)^tr^ and includes the following *n* = 9 transitions, each with its corresponding shift vectors

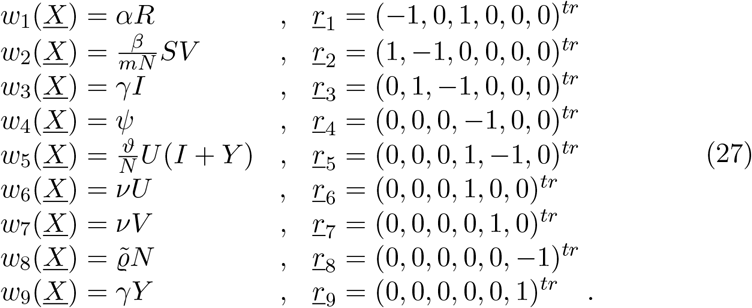

Similarly to the deterministic approach, the dynamics of imported cases *Y*, can be described on its own by the following master equation

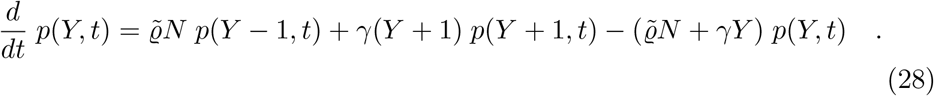

For the cumulative number of imported cases *C*_*Y*_, using the entry transitions of the stochastic dynamics of *Y*, we have the master equation given by

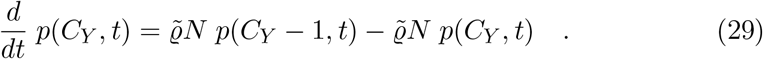

This represents a simple Poisson process with a known distribution in time evolution [47, 19].

The analytical solution to the master equation (29), given the initial condition *C*_*Y*_ (*t*_0_) at time *t*_0_, is described by the conditional probability

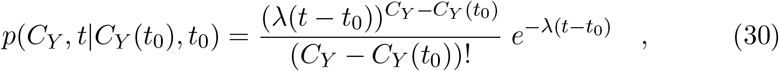

where 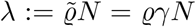 is constant. This represents a Poisson distribution for each time point *t*.

Figure 3(a) illustrates the differences between the mean field approximation and a stochastic realization for imported cases. We observe that, in contrast to the mean field approximation (in black), where the solution converges to a constant infected population, on average less than one infected case, the stochastic realization of imported cases (in magenta) can exhibit time intervals with no cases and others with multiple cases. This pattern is also observed in the cumulative imported cases, as shown in Figure 3(b).

**Figure 3:**
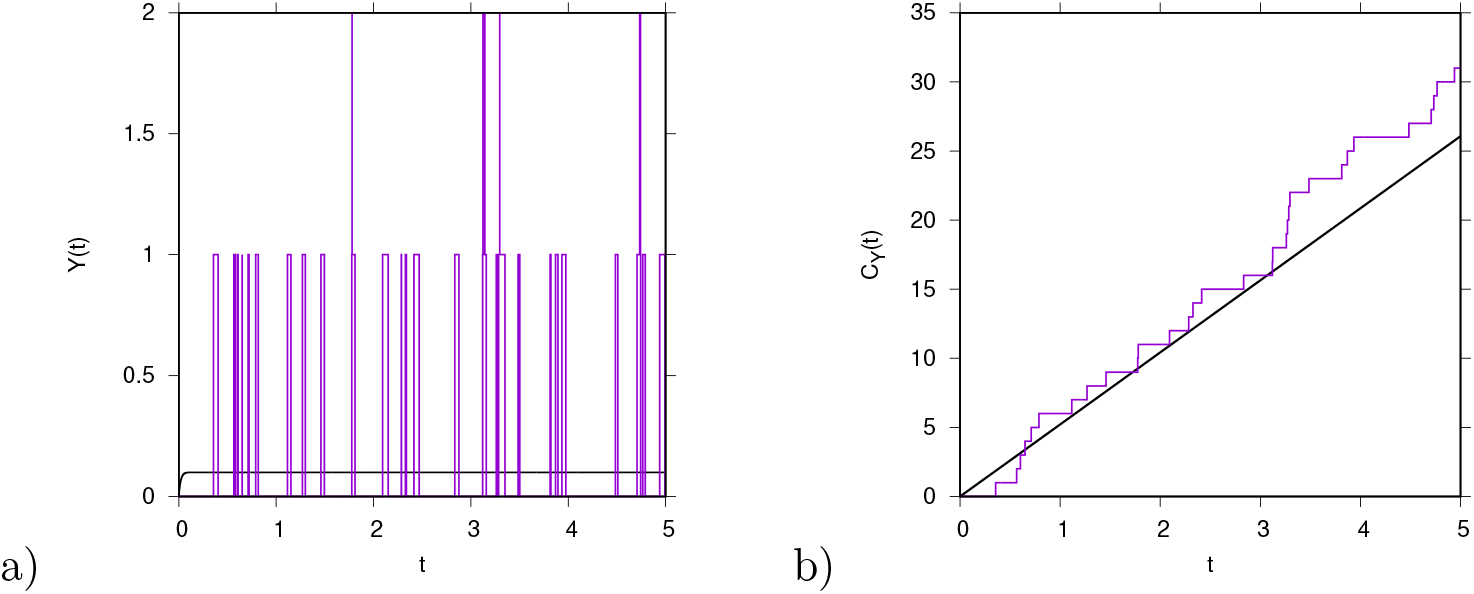
One stochastic realization (in magenta) and the mean field solution (in black) of (a) imported cases *Y* (*t*), and (b) cumulative imported cases *C*_*Y*_ (*t*). For *N* = 10^4^ and ϱ = 10^−5^, the parameter values used in this simulation are listed in Table 1.

Motivated by stochastic modeling, we apply statistical methods to model imported cases and integrate these models with our risk estimator. Let *τ* denote the waiting time between two consecutive imported cases, *C*_*Y*_ (*t*_*n*−1_) to *C*_*Y*_ (*t*_*n*_). The probability distribution of *τ* is given by

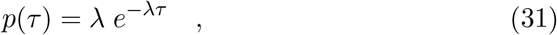

where *λ* := ϱ*γN* is the rate parameter. This distribution allows us to analyze the statistical characteristics of imported case notifications based on the waiting times between their occurrences.

#### 2.6.1 Parameter estimation from waiting time distribution

Given the recorded imported cases and the time intervals between consecutive cases, represented as the data vector <u>*τ*</u> = (*τ*_1_, *τ*_2_, …, *τ*_*n*_), we estimate the parameter *λ* using the log-likelihood method.

The likelihood function for the waiting times is given by

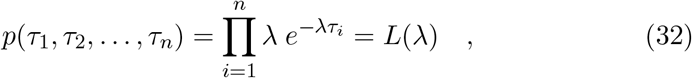

where the log-likelihood function is maximized to determine the maximum likelihood estimator

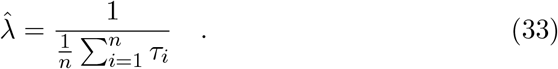

From the given data, we order the waiting times to produce the observed distribution function (shown in magenta in Figure 4). This is then compared to the cumulative distribution function

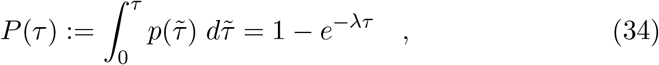

using both the known parameter *λ* = ϱ*γN* (represented by the black line in Figure 4) and the maximum likelihood estimator 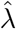 (middle green line in Figure 4).

**Figure 4:**
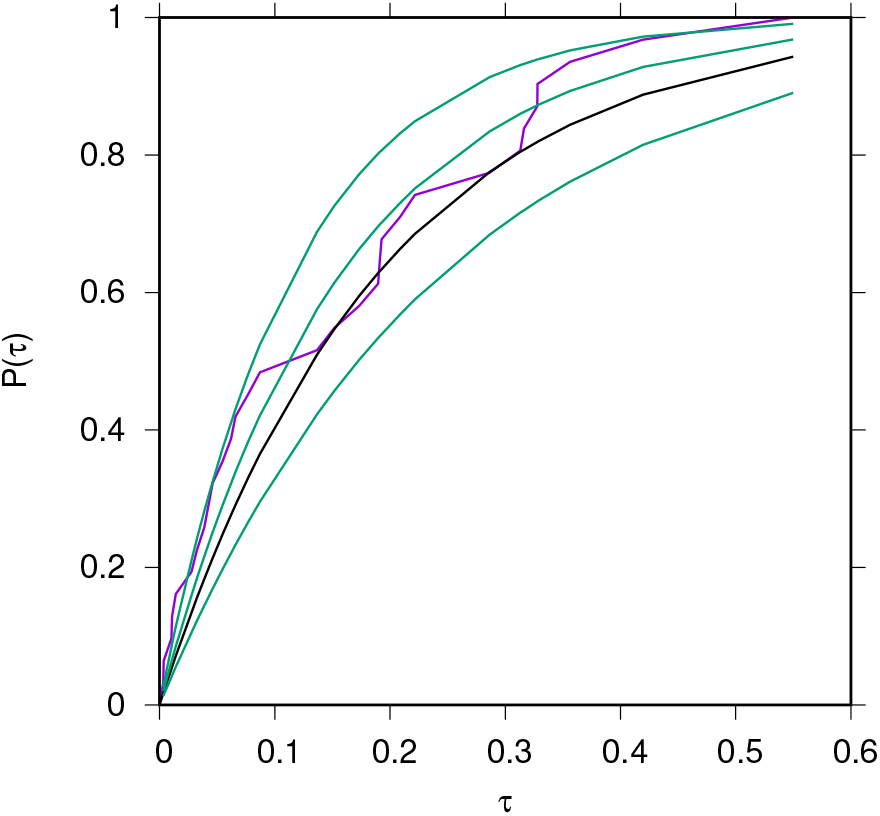
The empirical cumulative distribution function is shown in magenta, while the maximum likelihood estimate and its 95% confidence intervals are represented by the green lines. The theoretical cumulative distribution, represented by a black line, is known in simulations but not in empirical data analysis. The numerical values are 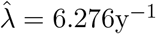 and ϱ*γN* = 5.214 y^−1^.

We then quantify the confidence interval of the estimation, acknowledging that simulated data may deviate from the true parameters. Once the best estimate of 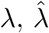, is determined, the expected number of imported cases can be expressed as

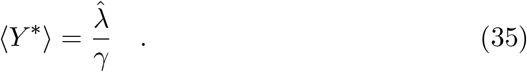

Although the risk estimator refers to average values, the model accounts for the intrinsic stochasticity of the time series.

#### 2.6.2 Confidence intervals from the likelihood function

Using the negative inverse Fisher matrix, which simplifies to a 1 × 1 matrix in the case of a single parameter, we can approximate the variance of a Gaussian distribution by examining the curvature of the log-likelihood function around its maximum. This variance is given by

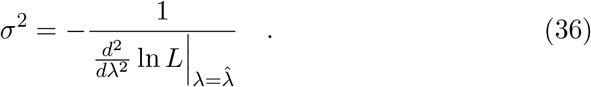

For the exponential waiting time likelihood

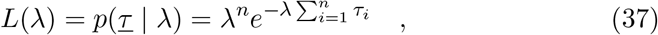

the variance simplifies to

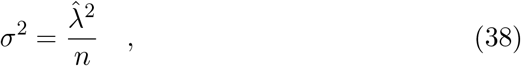

where *n* is the number of data points. This provides a 2*σ* confidence interval, which approximately covers 95% of the Gaussian distribution

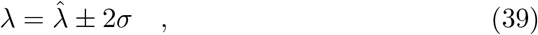

as shown in Figure 4).

The estimate of *λ* can be applied to the cumulative distribution function *P* (*τ*). In Figure 4, *P* (*τ*_*i*_) is obtained by ordering the *τ*_*i*_ values from smallest to largest (x-axis) and plotting 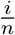 on the y-axis (in magenta), where the *τ*_*i*_ values are derived from simulations with *λ* = ϱ*γN*. The figure also includes the maximum likelihood estimate 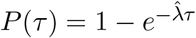 (middle green line) along with its 95% confidence intervals (upper and lower green lines). Additionally, the theoretical cumulative distribution function 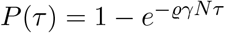 is shown (black line).

For a more thorough analysis of estimation uncertainties, Bayesian methods using conjugate priors, such as a Gamma distribution for the exponential case, could be used. The Bayesian posterior *p*(*λ* | *τ*) may provide asymmetric confidence intervals. For further details on these methods, see [47], as well as Appendices A and B for a more in-depth discussion in the present context.

Note that the current analysis of inter-event times is preliminary, providing a basic guide for stochastic simulations. As more data become available, the analysis can be refined and more sophisticated statistical methods can be applied.

#### 2.6.3 Derivation of the maxima likelihood estimate variance and theoretical background

For large data sets, the likelihood function approximates a Gaussian distribution around the maximum log-likelihood estimator. Specifically, the likelihood function *L*(*λ*) can be approximated by a Gaussian distribution *p*_*G*_(*λ*), given by

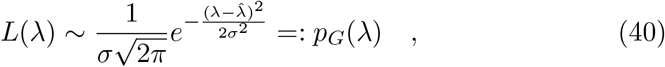

where 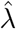 is the mean value and *σ*^2^ is the variance, determined by the curvature of the log-likelihood function around the maximum. This curvature is quantified by the second derivative of the log-likelihood function at the maximum, expressed as

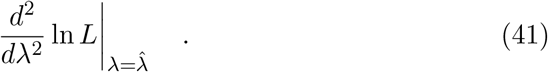

For the Gaussian approximation, the variance is

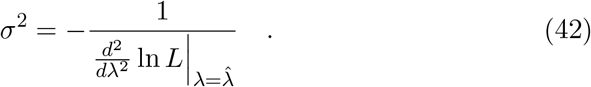

Applying this to the likelihood function of the exponential distribution

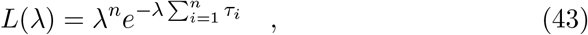

we obtain

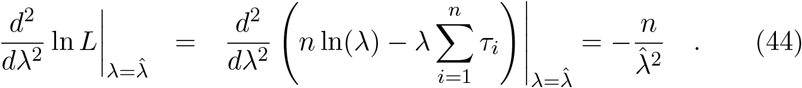

Thus, the variance is

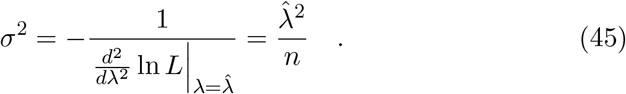

The standard deviation is 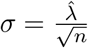, which can be used to determine the confidence interval for *λ* as 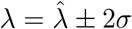. While this approach is practically useful, a more fundamental description and justification is provided in Appendix B through the Cramér-Rao consideration, which offers only an inequality as a lower bound for Equation (45).

## 3 Mosquito abundance and its impact on disease transmission

Analyzing mosquito abundance is crucial for predicting the transmission of mosquito-borne diseases, but it presents significant challenges. To address these complexities, a practical approach is to establish a threshold for mosquito abundance. Below this threshold, the likelihood of local disease transmission is minimal, while exceeding it significantly increases the risk of autochthonous cases.

### 3.1 Explicit calculation of Poisson process approximation for autochthonous cases

To model the dynamics of autochthonous infected cases, we start with the differential equation

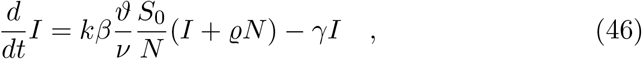

where 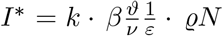, see Equation (23), and *ε* → *γ*. For small *k*, the equilibrium value *I*^*^ approaches zero, simplifying the dynamics.

The cumulative number of autochthonous cases *C*_*I*_ then follows

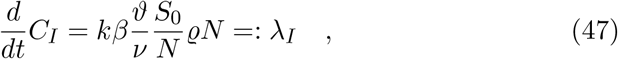

where *λ*_*I*_ represents the rate of new autochthonous cases.

Thus, the stochastic process for *C*_*I*_ can be approximated by a Poisson process. The master equation for *p*(*C*_*I*_, *t*) is

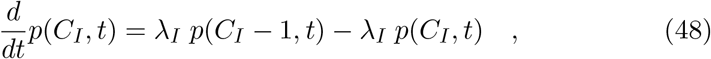

which has the Poisson distribution solution

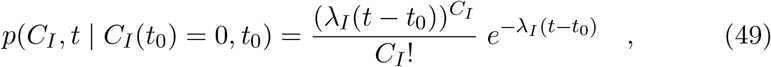

Where *C*_*I*_(*t*_0_) = 0 and *T* = *t* − *t*_0_ represents the time interval, similar to the analysis of imported cases, denoted by *C*_*Y*_.

The probability of having no autochthonous cases is

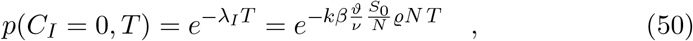

which approaches 1 as *k* becomes small.

The probability of having one or more autochthonous cases is

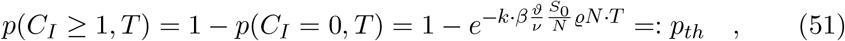

where *p*_*th*_ is a threshold probability. Solving for *k* yields

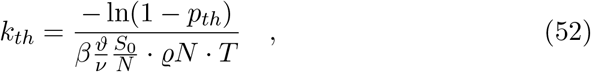

which gives the mosquito abundance *k*_*th*_ needed to achieve a certain probability *p*_*th*_ of having one or more autochthonous cases. This can be used for different values of *p*_*th*_, such as 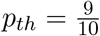 or 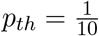, in subsequent analyses.

### 3.2 Analysing the mosquito abundance threshold

In this section, we analyze the bounds for relative mosquito abundance based on theoretical scenarios. We assume *S*_0_ = *N*, ϱ = 10^−5^, *N* = 10^4^, and *Y* ^*^ = 0.1. The threshold values for mosquito abundance *k*, for a given probability *p*_*th*_ are determined using Equation (52).

The relationship between imported cases and local transmission, emphasizing the role of mosquito abundance in determining the probability and timing of autochthonous cases, is illustrated in Figure 5. In a real-world scenario, this is evident in the comparison between the Basque Country, which has reported zero non-travel-related cases so far, and Italy, which had over 80 cases notified in 2023 [11], highlighting the different risks potentially associated with varying mosquito densities and environmental conditions.

**Figure 5:**
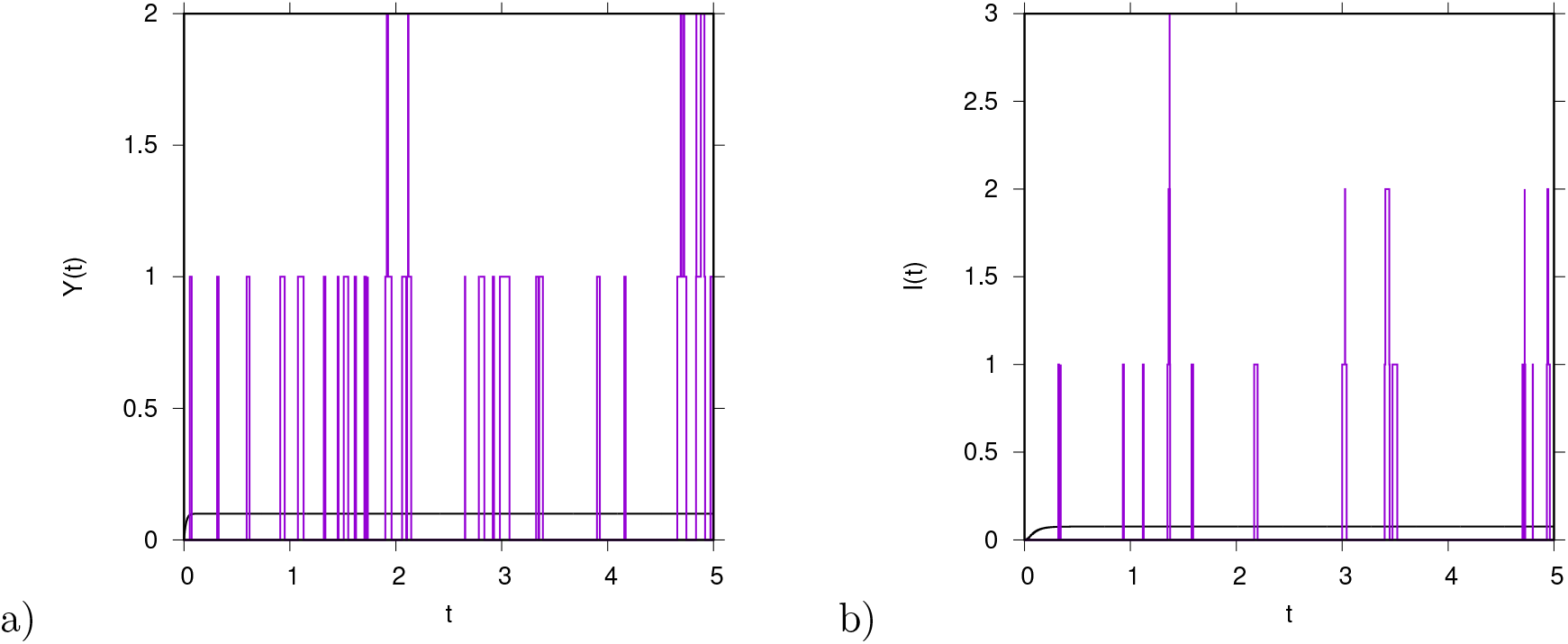
For mosquito abundance *k* = 30%, ϱ = 10^−5^, and *N* = 10^4^, in a) imported cases *Y* (*t*) and b) autochthonous cases *I*(*t*). Stochastic simulation is shown in magenta and its mean-field solution in black.

Considering a probability threshold of 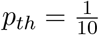, which means that in 1 out of 10 stochastic runs we expect at least one autochthonous case within the time interval *T* = 1 year, we obtain

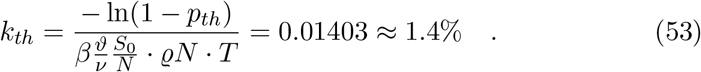

For a higher probability threshold of 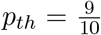, the value of *k*_*th*_ is approximately 31%. This indicates that we would nearly always observe at least one case in stochastic simulations within the first year.

Figure 5 illustrates the relationship between imported cases and local transmission, highlighting how mosquito abundance impacts the likelihood and timing of autochthonous cases to occur. For a mosquito abundance of *k* = 30%, Figure 5(a) shows the time series of imported cases *Y* (*t*), while Figure 5(b) presents the time series of autochthonous cases *I*(*t*). Two autochthonous cases occur in the first year (shown in magenta), but periods of up to a year with no autochthonous cases also occur, illustrating the stochastic nature of disease transmission even at relatively high mosquito abundance.

The Poisson distribution for the threshold value *k*_*th*_ = 0.3067 ≈ 30% is presented in Figure 6, showing the probability of no autochthonous cases being *p*(*C*_*I*_ = 0, *T*) = 0.1 and the probability of one or more cases being *p*_*th*_ = 0.9.

**Figure 6:**
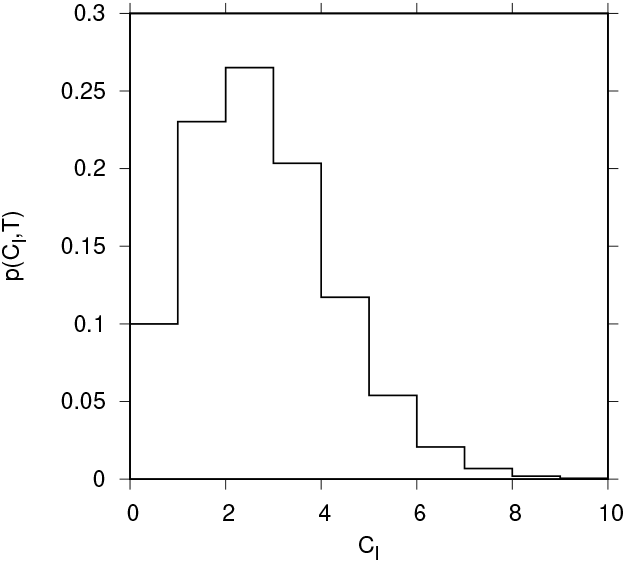
Poisson distribution for *k*_*th*_ = 0.3067 ≈ 30 %.

Conversely, for the threshold value *k*_*th*_ = 0.01403 ≈ 1.4%, Figure 7(a) demonstrates that the probability of observing zero autochthonous cases is *p*(*C*_*I*_ = 0, *T*) = 0.9, leading to *p*_*th*_ = 0.1 for one or more cases. Figure 7(b) indicates that with this threshold, we nearly always observe no autochthonous cases within the first year and rarely any cases in subsequent years.

**Figure 7:**
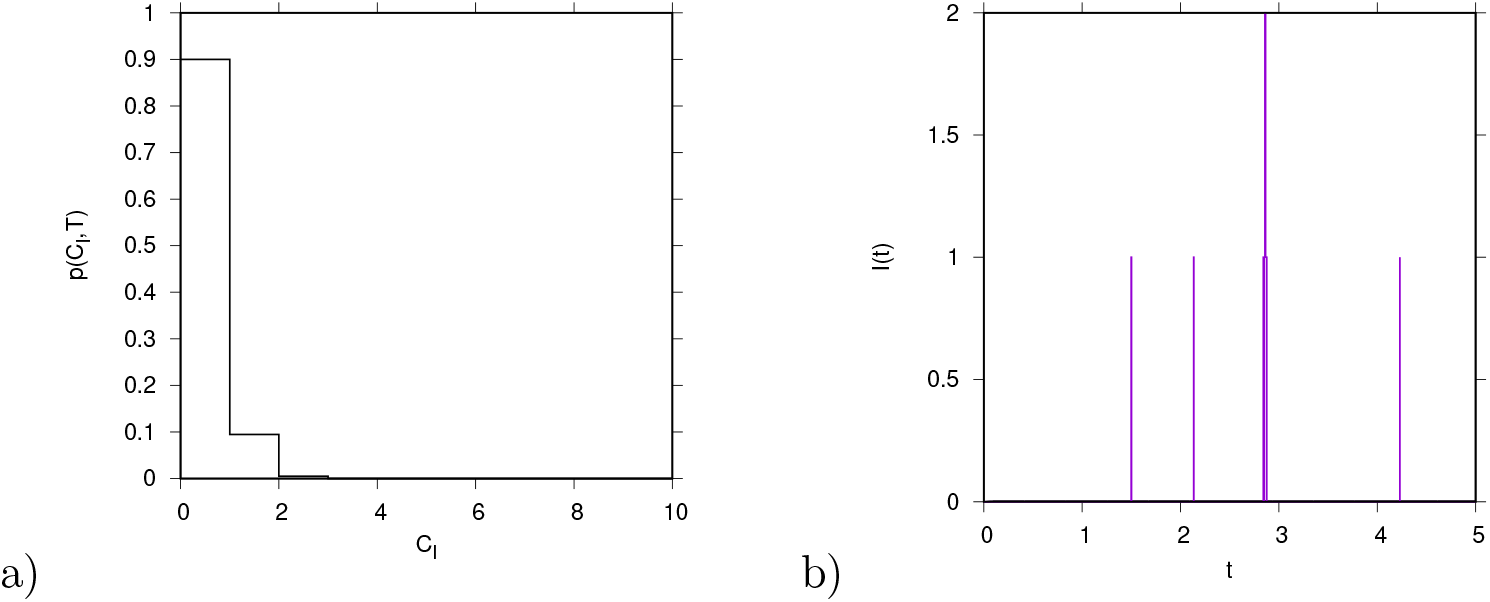
a) Poisson distribution for *k*_*th*_ = 0.01403 ≈ 1.4%. b) Time series of autochthonous cases *I*(*t*), for a mosquito abundance of *k* = 2%.

While the Poisson process approximation for the autochthonous cases *I*(*t*), especially for small mosquito abundance *k*, provides an indication of the expected frequency and duration without cases, the full stochastic model simulations reveal more clustering of cases. For example, Figure 5(b) shows up to three cases occurring simultaneously, and Figure 7(b) shows two cases at the same time. This clustering may be influenced by state-dependent factors in the stochastic variables *w*_*j*_ that are not captured by the basic Poisson approximation.

### 3.3 Analysis of the distance from exponential growth *ε*(*k*)

For small mosquito abundance *k*, the term *ε*(*k*), defined in Section 2.4, can be approximated by the recovery rate *γ*. This approximation is illustrated in Figure 8, where *k* values are shown on a logarithmic scale (base 10). For instance, for *k* = 1%, the approximation *ε* ≈ *γ* is already quite accurate, with *γ* ≈ 52 y^−1^.

**Figure 8:**
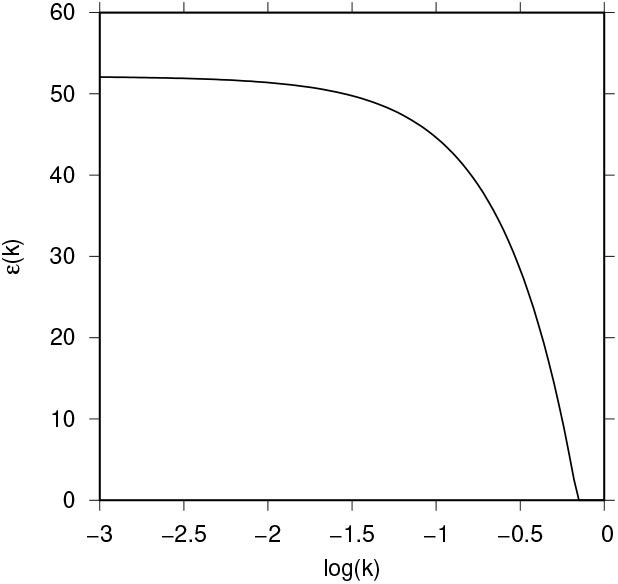
Values of *ε*(*k*), the distance to the threshold of exponential growth, converge towards *γ* = 52y^−1^ as the relative mosquito abundance *k*, decreases.

In an invasion scenario with relatively low mosquito abundance and no observed autochthonous cases, the expected number of autochthonous cases ⟨*I*⟩ can be quantified using the simplified expression

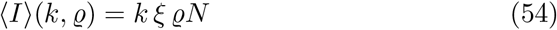

where 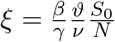 is constant. For a naive, completely susceptible population,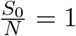. Thus, *ξ* can be compared to the basic reproduction number typically expected in vector-borne disease models. Consequently, our risk estimator is given by Equation (54).

It is important to note that, due to the presence of several imported cases over time, we might still expect occasional autochthonous cases, even if they occur infrequently. Additionally, observed increases in mosquito abundance over the years [11] could impact this risk.

The primary focus of the risk measure is to determine the probability of observing the first autochthonous case in a non-endemic area. Although the calculated expectation ⟨*I*⟩ is well below one (see Figure 1), stochastic simulations can provide insights into when the first cases might occur, depending on the levels of imported cases and mosquito abundance. We will analyze scenarios ranging from low to intermediate mosquito abundance and examine conditions where the approximation *ε* ≈ *γ* no longer holds. In such cases, the non-linearity of *ε*(*k*) can lead to increasing clusters of outbreaks until the threshold for exponential growth and self-sustained transmissibility is reached.

### 3.4 The risk of outbreaks in increasing mosquito abundance scenarios

In this section, we analyze scenarios where mosquito abundance increases beyond the simplest approximation explored previously. To analyze deviations from the simplest linear relationship of the expected number of autochthonous infections ⟨*I*⟩_*_ with respect to mosquito abundance *k*, we need to consider how the effective transmission rate *ε*(*k*) changes as *k* increases.

#### 3.4.1 Deviations from the simplest linear relation of expected autochthonous infected on mosquito abundance

The simplest risk estimator for the expected number of autochthonous infections ⟨*I*⟩_*_ is given by

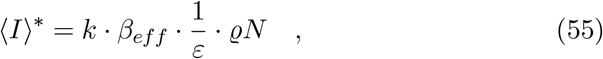

where *ε* is the distance to the threshold of exponential growth and is given by

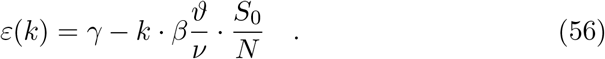

In the initial phases of outbreaks, where *S*_0_ ≈ *N*, and as long as *ε >* 0, the expression *ε*(*k*) → *γ* as *k* → 0 indicates that the distance to the threshold of exponential growth approaches the recovery rate *γ*.

However, deviations from the simplest linear relationship become significant as mosquito abundance *k* approaches the critical threshold *k*_*c*_. In this regime, the linear approximation of 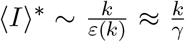 fails to hold, and the expected number of autochthonous cases can increase very rapidly.

As shown in Figure 9, the relationship between ⟨*I*⟩_*_ ∼ *k/ε*(*k*) and mosquito abundance reveals that for small *k* the expression *k/ε*(*k*) approximately behaves linearly as *k/γ*. This is expected since *ε*(*k*) ≈ *γ* for small values of *k*. Nevertheless, close to the critical threshold *k*_*c*_ there is a significant non-linear growth in ⟨*I*⟩_*_. As *k* approaches *k*_*c*_, *ε*(*k*) decreases rapidly, causing ⟨*I*⟩_*_ to rise sharply. This non-linearity is due to the rapid approach of *ε*(*k*) towards zero, leading to a divergence in ⟨*I*⟩_*_. These findings highlight the importance of accounting for the non-linearity of *ε*(*k*) in disease transmission models and emphasize the need for careful empirical observation of mosquito abundances, as they may increase from year to year.

**Figure 9:**
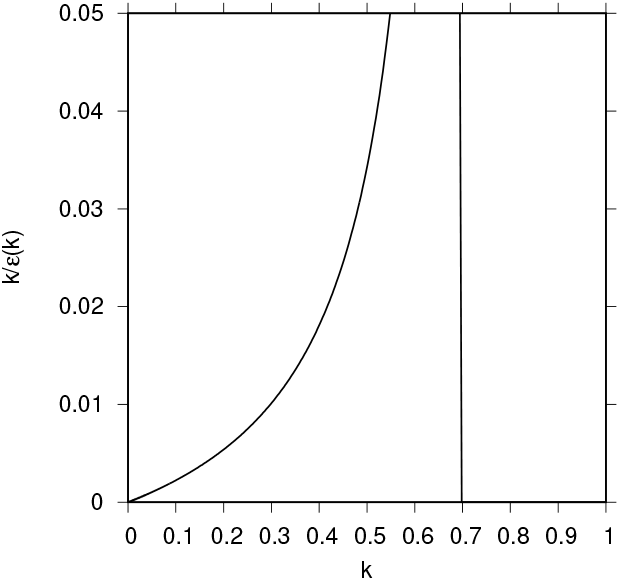
Graphical representation of the expression 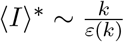 as a function of the relative mosquito abundance *k* (bent curve). The vertical line represents the critical value *k*_*c*_.

#### 3.4.2 Temporal dynamics and scaling close to the epidemiological threshold

The dynamics described in Equation (21) for the sub-critical regime reads

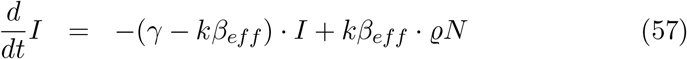

and can be solved, yielding its time-dependent solution given by

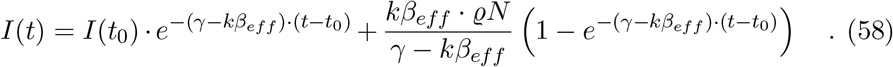

When the mosquito abundance *k* reaches its threshold value *k*_*c*_, given by *γ* − *k*_*c*_ · *β*_*eff*_ = 0, the time-dependent solution is

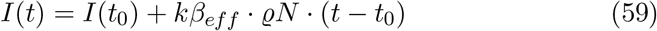

i.e. with the distance to threshold *ε* = *γ* − *kβ*_*eff*_ = 0. This implies that close to the threshold, the total number of infected individuals becomes asymptotically self-similar, i.e., a homogeneous function of the general form

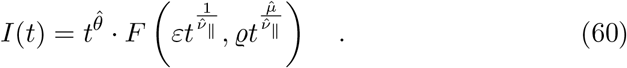

The critical exponents 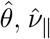, and 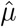 characterize the self-similar, scale-invariant behavior of the infection spread near the critical threshold, governing the growth, time evolution, and density fluctuations of the infected population [48].

Rescaling in terms of the parameter *ε*, we have

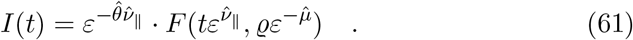

In the mean-field regime, i.e., no spatial effects, the exponents are 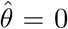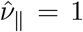, and 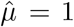 (see [48] for further details on general scaling theory around critical thresholds).

For empirical epidemiological systems, this results in distributions of avalanches with power-law tails, leading to larger numbers of autochthonous cases more frequently than expected, for example, in classical Gaussian distributions.

Thus, from a management point of view, once mosquito abundance increases and leads to larger clustered outbreaks, even small additional increases in mosquito numbers will result in significantly larger outbreaks of autochthonous cases.

## 4 Applying the model to Basque Country data

The Basque Country is an autonomous region in northern Spain with approximately 2.2 million inhabitants. In this region, the Public Health Epidemiological Unit, in collaboration with NEIKER - the Basque Institute for Agricultural Research and Development - monitors records of both imported disease cases and the distribution and establishment of disease vectors. Although the Basque Country is considered a non-endemic area for tropical mosquito-borne diseases, it has established populations of *Aedes* mosquitoes (with increasing distribution) and frequently reports viremic imported cases of dengue, Zika, and Chikungunya. This makes it an ideal setting for this study.

### 4.1 Imported cases of *Aedes* mosquito-borne diseases in the Basque Country in 2019 (Pre-COVID) and 2022 (Post-COVID)

The raw empirical data of cumulative imported cases of dengue, Zika and chikungunya in the Basque Country for the years of 2019 and 2022, provided by the Health Basque Department, is illustrated in Figure 10.

**Figure 10:**
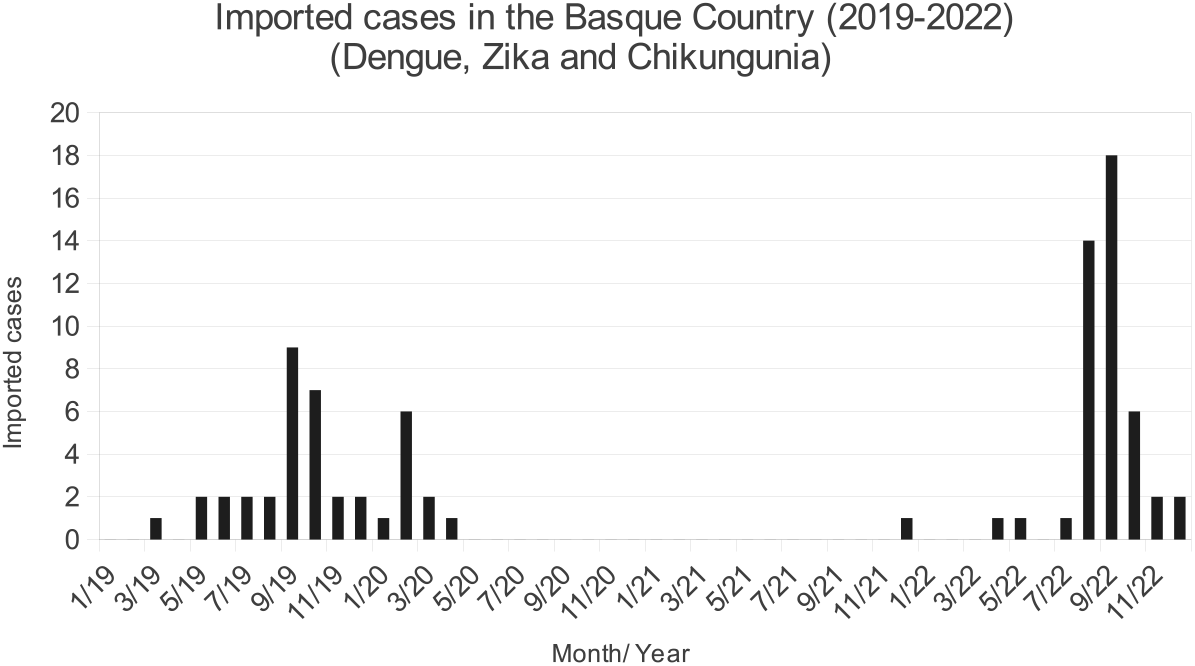
Epidemiological records of imported disease cases in the Basque Country from 2019 to 2022.

The data reveals a steady registration of imported cases throughout the year, with a notable increase during the summer months. This seasonal surge coincides with the period of peak mosquito activity, which can contribute to higher disease transmission rates. Note that for the years of 2020 and 2021, very few imported cases were recorded since travel restrictions were in place due to the COVID-19 pandemic.

#### 4.1.1 Estimation of imported cases

Applying the statistical analysis described in Section 2.6.2, we obtain the inter-event cumulative distribution of imported cases, and the cumulative counts of imported cases of dengue, chikungunya, and Zika, i.e. *Aedes* mosquito-transmitted diseases notified in the Basque Country for the years 2019. For 2022, there were no significant differences in the number of imported cases compared to 2019; the numbers remained relatively similar, although travel activity was higher in the latter half of the year. The primary difference was the increase in mosquito abundance, as illustrated in Figure 12.

For 2019, Figure 11 (a) shows the inter-event cumulative distribution of imported cases *P* (*τ*) for dengue, chikungunya, and Zika in the Basque Country (in blue), alongside the inter-event rate *λ* = (0.085 ± 0.031) d^−1^ (in green). Figure 11 (b) shows the cumulative number of imported cases *C*_*Y*_ (*t*) for dengue, chikungunya, and Zika (in blue) and the estimated mean number of cumulative imported cases 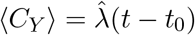 (in green).

**Figure 11:**
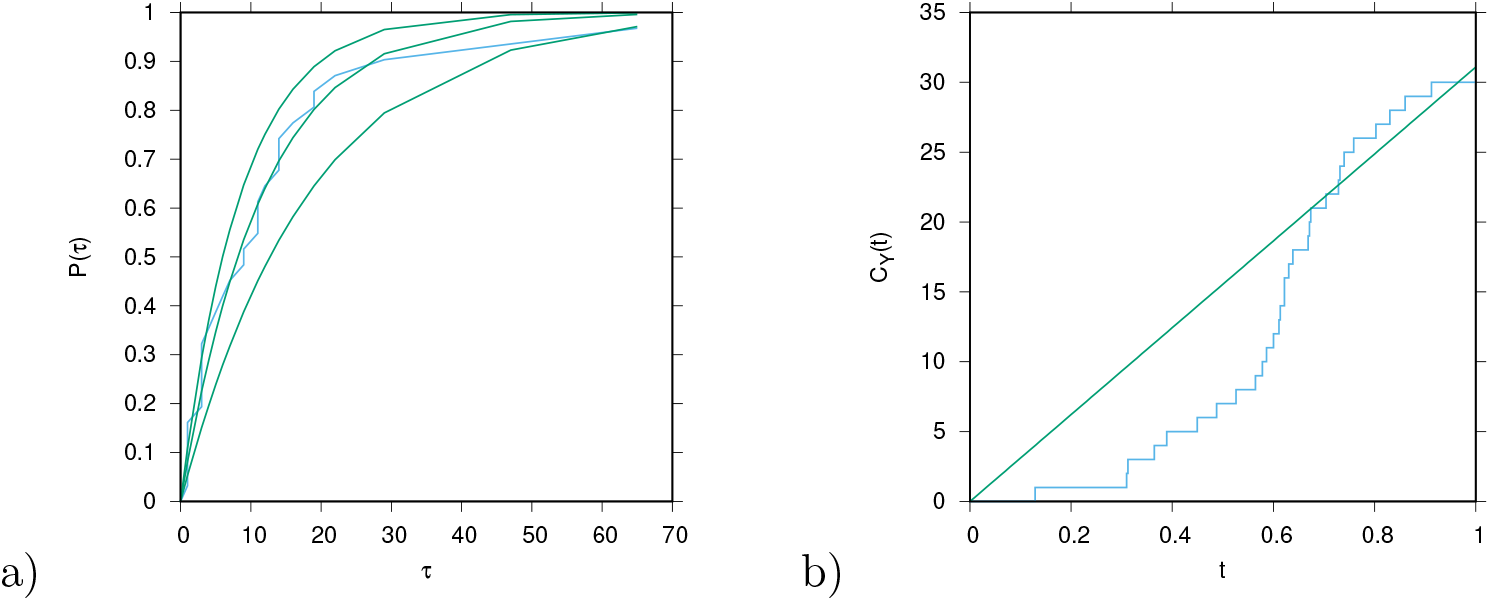
For the year of 2019, in (a) the inter-event cumulative distribution of imported cases *P* (*τ*) (in blue), alongside the inter-event statistics (in green). In (b) the cumulative number of imported cases *C*_*Y*_ (*t*) (in blue), and the estimated mean number of cumulative imported cases 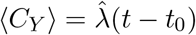 (in green). The inter-event times *τ*_*i*_ include the first and the last day of the year.

**Figure 12:**
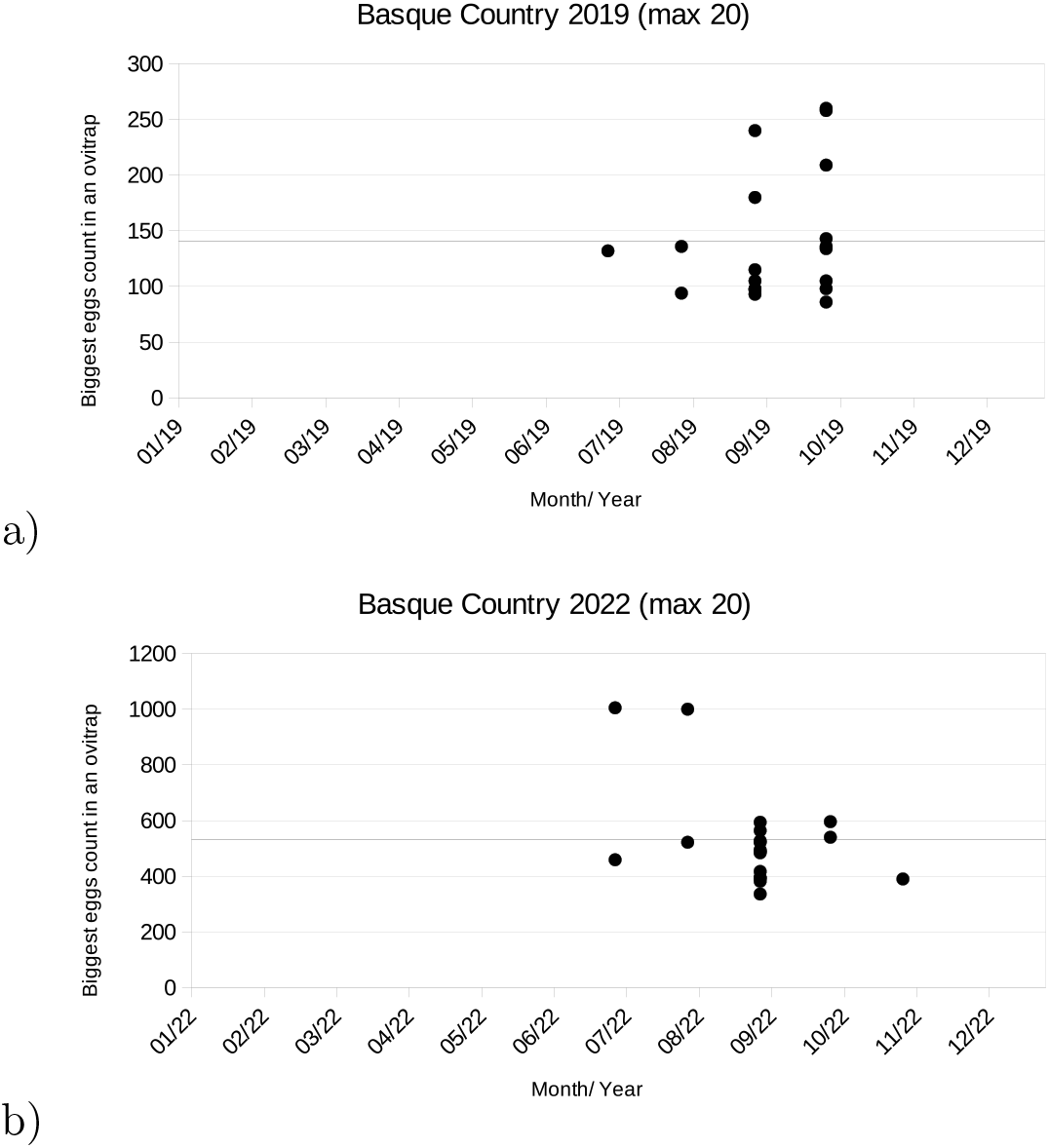
The largest 20 eggs counts in Basque Country in a) 2019, and b) 2022.

The estimation of imported cases in the Basque Country is derived from the data on imported cases using maximum likelihood estimators. The estimation of the parameter *λ* is given by Equation (33). The confidence interval for *λ* is calculated as 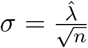. For the Basque Country in 2019, this yields

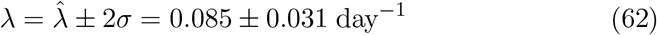

Additionally, data from the post-COVID-19 pandemic period in 2022 provides an estimated mean number of imported cases

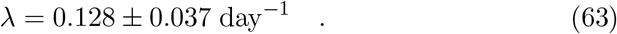

This indicates some increase in the number of imported cases in 2022. Variations in the risk of autochthonous cases across different years are evident, with a clear upward trend. Differences in trends may also be observed at different spatial scales, such as by province, though these differences are generally not substantial, especially if confidence intervals overlap. Furthermore, variations over the years show lower risks before August and after September, with increased risks during the summer months, aligning with the period of higher mosquito activity.

The expected number of imported cases ⟨*Y* ⟩_*_ is related to the Poisson rate *λ* as follows

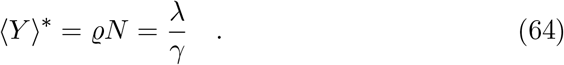

As an example, using the results for 2019 in the Basque Country, where *λ* = 0.1, we obtain

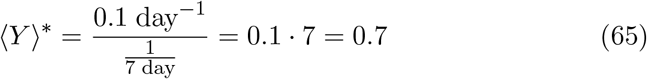

This initial statistical analysis provides insights into the trends of imported cases in the Basque Country, offering preliminary estimates of parameters critical for assessing the risk of mosquito-borne diseases in the region.

To further refine this analysis, a potential improvement would involve seasonal breakdowns, such as spring, summer, and autumn, to capture variations in case numbers. However, this approach faces challenges, including a smaller dataset for each season and the lack of robust patterns due to significant regional variations. Additionally, the sparse data for 2020 and 2021, due to travel restrictions from the COVID-19 pandemic, limits the analysis. Future data may reveal a return to more typical patterns and provide clearer trends.

### 4.12 Mosquito abundance data in the Basque Country

In the Basque Country, the Basque Institute for Agricultural Research and Development (NEIKER) has been monitoring mosquito breeding sites and recording egg count data across various localities for over a decade [49], providing increasingly detailed insights into mosquito population trends.

The data show a significant increase in egg counts from ovitraps in recent years. Notably, the counts in the most populated traps have reached levels comparable to those in endemic regions, with maximum counts nearing 1, 000 eggs. However, since the ovitraps are not yet standardized across temporal and spatial scales, comparing mosquito abundance between invasion scenarios and endemic areas remains challenging.

Given the increasing number of traps deployed each season, this analysis focuses on the sum of the highest 20 egg counts as a preliminary indicator of mosquito abundance in the Basque Country for 2019 and 2022.

Figure 12 shows the 20 highest egg counts in the Basque Country. There is a noticeable difference between 2019 (pre-COVID-19 pandemic) and 2022 (post-COVID-19 pandemic). In 2022, the maximum egg counts reached 1000 eggs, with a mean of approximately 530, while in 2019, the maximum count was only a quarter of that, with a mean around 140 eggs.

Since no autochthonous cases of *Aedes* mosquito-borne diseases have yet been detected in the region, these egg counts serve as initial indicators of mosquito abundance. These results can be applied to calculate the relative risk of disease transmission, considering the presence of imported cases and mosquito levels in the Basque Country.

#### 4.1.3 Risk of autochthonous cases in the Basque Country

The expected risk of autochthonous cases ⟨*I*⟩_*_ is given by

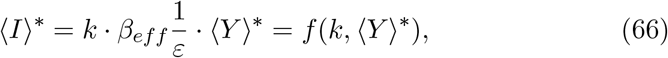

and for small mosquito abundance *k*, where *ε*(*k*) ≈ *γ*, this simplifies to

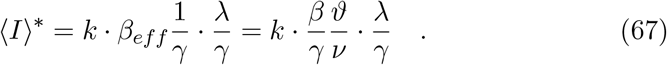

Here, 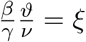 is a factor defined from basic information in endemic countries. As discussed in Section 2, we use 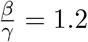 and 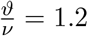, giving 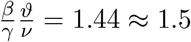 These values can be adjusted based on updated information for different vector-borne diseases.

The expected number of autochthonous cases in the Basque Country, based on mosquito abundance in 2019, can be estimated as ⟨*I*⟩_*_ = 0.0671, and in 2022 as ⟨*I*⟩_*_ = 0.5602, suggesting a potential rise in the number of cases. Although these estimates may indicate fewer than one person affected, stochastic simulations suggest that clusters of infected cases could emerge in the coming years.

Using this approach, the risk can be calculated on a smaller spatial scale, such as at the provincial and municipal levels, as more data become available. Additionally, risk maps with finer spatial resolution, such as at the municipality level, can be generated. These maps would consider imported cases, mosquito abundance, and the combined risk of autochthonous cases, providing a more detailed risk assessment.

## 5 Discussion and conclusion

Driven by climate change and increased global travel, mosquito-borne diseases like dengue, Zika, and chikungunya, once confined to subtropical and tropical regions, are now being reported in non-endemic areas. Although most cases originate from infected travelers - referred to as imported cases - they can trigger unexpected autochthonous outbreaks in regions with competent mosquito vectors. This has been evident recently, with non-travel-related dengue cases reported in France and Italy, as noted by the European Center for Disease Prevention and Control [11].

In endemic areas, mathematical models have been developed to estimate disease outbreak risks, with the Basic Reproduction Number (*R*_0_) being widely used to assess the likelihood and intensity of outbreaks occurring throughout the year. This metric, which represents the average number of secondary cases produced by an infected individual in a completely susceptible population, is crucial for understanding the dynamics of disease spread in regions where transmission is ongoing. However, in non-endemic areas where no local cases have been recorded, *R*_0_ is typically less than 1 (*R*_0_ *<* 1), indicating that the disease is not currently spreading within the local population. As a result, *R*_0_ is not a useful measure for assessing outbreak risks in these regions. Instead, alternative models and metrics must be used to account for the role of imported cases and the presence of competent mosquito vectors, which can introduce and potentially amplify disease transmission in previously unaffected areas.

In this study, a dynamic vector-host model is developed to assess the risk of autochthonous cases driven by imported infections in non-endemic regions. Using stochastic methods, the model analyzes waiting times for the first local cases and inter-event statistics of imported cases, with a specific application to the Basque Country. This approach incorporates imported cases and the presence of mosquito populations, relative to those in endemic regions, to estimate the risk of mosquito-borne disease outbreaks.

Stochastic simulations reveal that even in non-endemic regions with moderate mosquito abundance - such as 40% of that found in endemic areas - there is a potential for isolated autochthonous cases driven by imported infections, as well as the possibility of outbreak clusters. These simulations indicate that the dynamics of mosquito-borne disease transmission can be significantly influenced by the presence and density of mosquito populations, even at lower levels. By estimating the expected number of infected cases and assessing the risk of autochthonous infections based on imported case data and mosquito abundance, the model provides a robust tool for predicting and managing mosquito-borne disease risks in non-endemic areas. This approach allows public health authorities to anticipate potential outbreaks and implement targeted interventions to mitigate the risk of disease spread. By considering the Basque Country - a region in Spain with documented imported cases but no local cases to date - as a case study, we apply our method to assess the risk of an autochthonous disease outbreak. Essential data for this analysis include mosquito egg counts recorded by NEIKER across various localities and epidemiological information on viremic imported cases provided by the Epidemiology Unit of the Public Health Department of the Basque Country.

Using a Poisson process, we analytically computed the trend of imported cases and applied this method to data from the Basque Country for 2019 and 2022. We also calculated the confidence intervals for the cumulative curve of imported cases via likelihood functions. It is important to note that for the years 2020 and 2021, very few or no imported cases were recorded due to travel restrictions imposed by the COVID-19 pandemic, resulting in insufficient data to apply our method effectively. Our analysis demonstrates that mosquito population abundance significantly impacts the likelihood and timing of autochthonous cases. This explains why autochthonous cases have been identified in countries like France and Italy but not yet in other European regions, such as the Basque Country. The findings underscore the critical importance of considering non-linear dynamics in disease transmission models, particularly near critical thresholds of mosquito abundance, and call for continued research into seasonality and other factors influencing mosquito abundance.

By coupling mosquito abundance calculations with the estimated mean number of imported cases in 2019, we apply the risk estimator developed through the stochastic framework. The expected number of autochthonous cases in the Basque Country rose from ⟨*I*⟩ = 0.0671 in 2019 to ⟨*I*⟩ = 0.5602 in 2022, and this increasing trend is anticipated to continue in the coming years. Although these estimates suggest fewer than one case on average, stochastic simulations indicate that clusters of infected cases could emerge in the coming years.

Our approach allows for risk calculation on a smaller spatial scale, such as at provincial and municipal levels, as more data becomes available. The spatial distribution of imported cases versus the measurements points of ovitraps pose some challenges on spatial correlation of the risks, however it facilitates the creation of risk maps with finer spatial resolution, such as at the municipality level, which will be investigated in future research, providing a more detailed risk assessment for public health authorities.

## Data Availability

All data produced in the present work are contained in the manuscript

## Acknowledgments

This work is also supported by the ARBOSKADI project for monitoring vector-borne diseases in the Basque Country, Euskadi. We wish to extend our acknowledgments to Jesús Ángel Ocio Armentia, Oscar Goñi Laguardia and Ana Ramírez de La Peciña Pérez, Dirección de Salud Pública for their fruitful discussions, and to Madalen Oribe Amores, Unidad de Vigilancia Epidemiológica de Bizkaia, for her cooperation in providing the requested epidemiological data that was essential for carrying out this research.

M.A. acknowledges the financial support by the Ministerio de Ciência e Innovacion (MICINN) of the Spanish Government through the Ramon y Cajal grant RYC2021-031380-I. This research is also supported by the Basque Government through the “Mathematical Modeling Applied to Health” (BMTF) Project, BERC 2022-2025 program and by the Spanish Ministry of Sciences, Innovation and Universities: BCAM Severo Ochoa accreditation CEX2021-001142-S / MICIN / AEI / 10.13039/501100011033.

## Appendices

### A Analysis of mean and variance via hypothetical ensembles of data

In this section, we summarize both the frequentist and Bayesian approaches to parameter estimation, along with their theoretical foundations.

In the previous analysis, we constructed the maximum likelihood estimator (MLE) and assessed its variance using frequentist methods. The variance of the MLE was estimated as 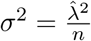, which was derived from the curvature of the likelihood function.

To theoretically justify this result, we consider the likelihood function *L*(*λ*) = *p*(<u>*τ*</u> | *λ*), which describes the probability of observing data ensembles given a particular model. The Gaussian approximation of the likelihood function provides a good fit for the distribution of estimators 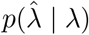. This fit can be validated through simulations that evaluate multiple data sets and examine the resulting histogram of estimators 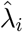. Although an exact analytical solution for the ensemble variance 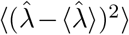 is not feasible, it can be approximated using the variance expression obtained from the negative inverse Fisher matrix [50]

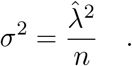

This expression is consistent with the Cramér-Rao inequality [51], which provides a lower bound for the variance of *λ* given by *λ*^2^*/n*. The Cramér-Rao inequality involves the true but unknown parameter *λ*, while our heuristic method uses the estimator 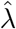. The complete form of the inequality also accounts for any bias in the estimator, resulting in

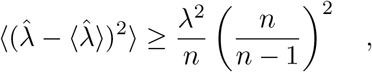

where the impact of bias diminishes quickly. Detailed calculations related to the Fisher information and the Cramér-Rao inequality can be found in Appendix B.

The likelihood function *L*(*λ*) = *p*(*τ* _*D*_ | *λ*) from a single empirical data set <u>*τ*</u>_*D*_ approximates the functional form of the distribution 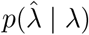 obtained by marginalizing over all possible hypothetical data sets *τ*, even if the estimator 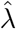 deviates from the original parameter *λ*.

In contrast, Bayesian analysis updates prior beliefs about the parameter *λ* using the empirical likelihood function from a single data set. The prior distribution *p*(*λ*), specified before observing the data, is updated to form the posterior distribution *p*(*λ* | <u>*τ*</u>_*D*_). This posterior provides estimates and confidence intervals and can be applied to the empirical data of imported cases in the Basque Country, as demonstrated in the analysis for the Basque Country. Due to the very limited data, the maximum likelihood method of frequentists provides reasonable information, while the more elaborate Bayesian approach might not refine the results significantly in the present study. Hence, we focus on the frequentist framework here.

### B. Calculation of the lower bound of the ensemble variance of maximum likelihood estimators

#### B.1 Conditioned ensemble averages, the starting point of the analysis, to the variance inequality

Conditioned ensemble averages serve as the starting point for analyzing the variance inequality. For the ensemble mean of estimators, we have

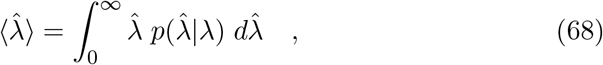

where the probability 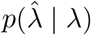 of estimators 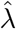, given a stochastic model with parameter *λ*, can be expressed by marginalizing over ensembles of data sets *τ* obtained from the likelihood function

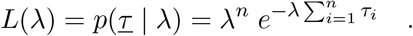

The likelihood function *L* gives the probability for the data given an underlying model with parameter *λ*. These data determine the estimators 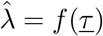 via the probability

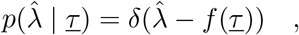

where 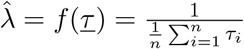, with the analytic functional form of the maximum likelihood estimator, giving

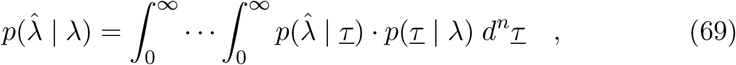

which essentially marginalizes via

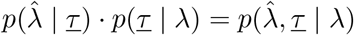

the joint probability 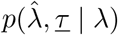 over *τ*. Hence, we have

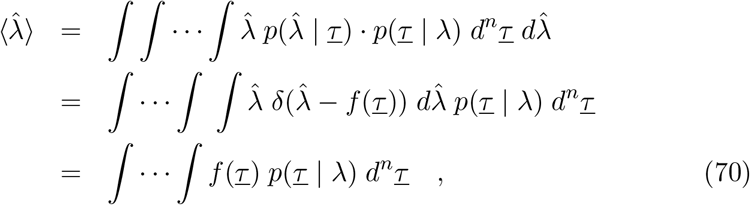

which gives, for the exponential distribution, after some calculations, the explicit result

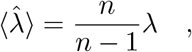

essentially via deriving and solving the ordinary differential equation

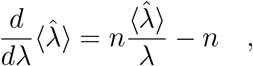

via the variable transformation 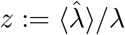, hence

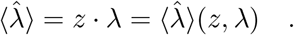

Then, the bias of the estimator is defined as the deviation of the ensemble mean of the estimator 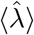 and the original parameter *λ*, that is

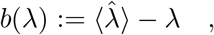

which is here in the case of the exponential distribution 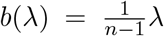, converging quickly to zero with increasing number *n* of data points. An analogous calculation for the variance of the estimator gives

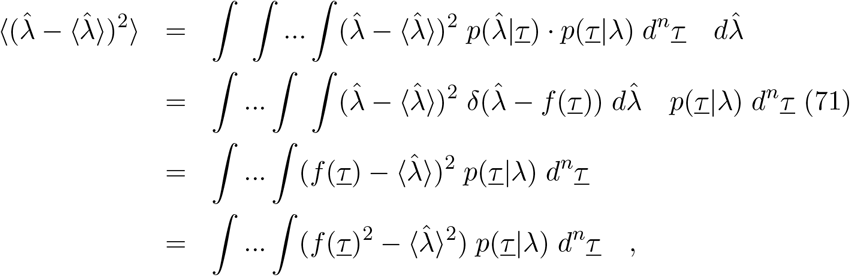

where in the last step we used in expectation 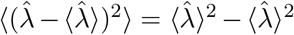. This can only be treated via an inequality giving a lower bound of the estimator variance.

In order to apply the Cauchy-Schwarz inequality, (which is in ordinary vector spaces relating the scalar product of two vectors to the Euclidian norm of the vectors as <u>*g*</u> · <u>*h*</u> ≤ ||<u>*g*</u>|| · ||<u>*h*</u>||), we first analyze

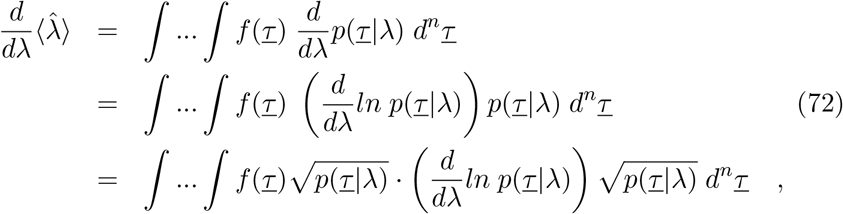

where in the last step the integrand is written as a product of two functions 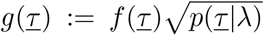 and 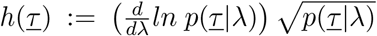 to give in the Cauchy-Schwarz inequality, now for integrals, the treatable expressions in form of expectation integrals.

After we considered 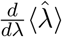 evaluating the definition of 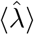 we now evaluate from the definition of the bias 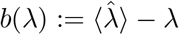, hence 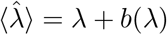 giving

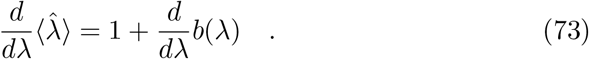

Applying, thus, the Cauchy-Schwarz inequality replacing the sums by integrals and the finite dimensional vectors by functions, we get

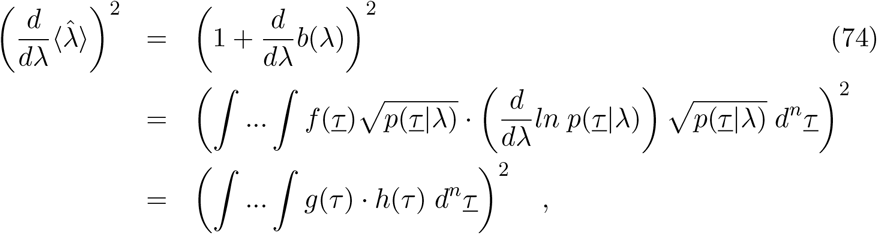

and applying the Cauchy-Schwarz inequality

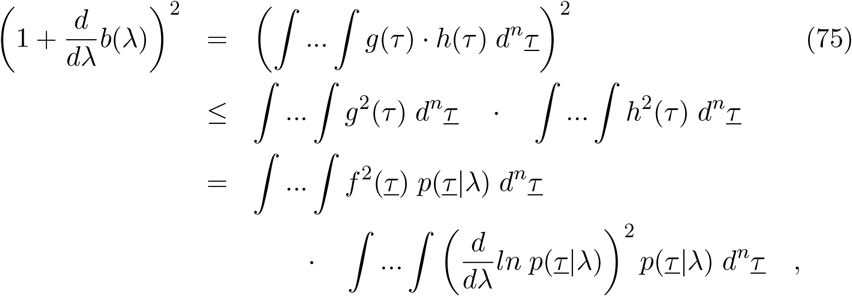

and hence, the inequality

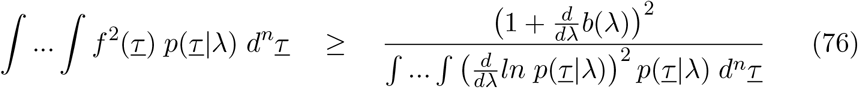

holds, which only needs little refinement for the final result.

For this, we consider now the normalization of the probability of the likelihood function ∫ … ∫ *p*(<u>*τ*</u> |*λ*) *d*^*n*^<u>*τ*</u> = 1 and its first two derivatives in respect to the parameter *λ*. We have

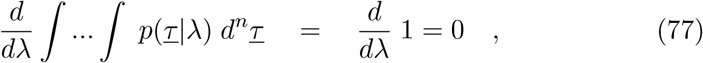

and hence

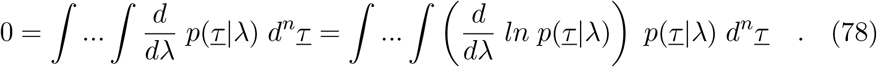

The zero can be multiplied by any function to still be zero, and any function not depending on the data <u>*τ*</u> can be taken into the integrals. Hence, for the function 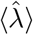 we obtain

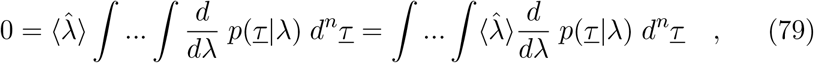

and with this we can rewrite Eq. (72) as follows

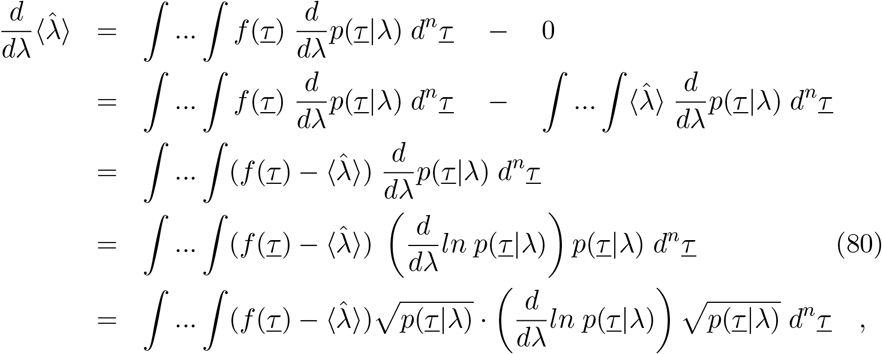

and redefining 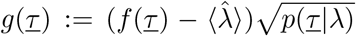 allows us, in all subsequent calculations above, to replace *f* (*τ*) by 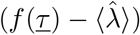. Hence, in Equation (76) we have now the complete variance on the left hand side of the inequality

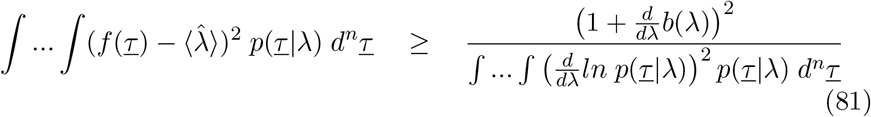

as desired.

The final step is to take the second derivative of the normalization of the likelihood function given by

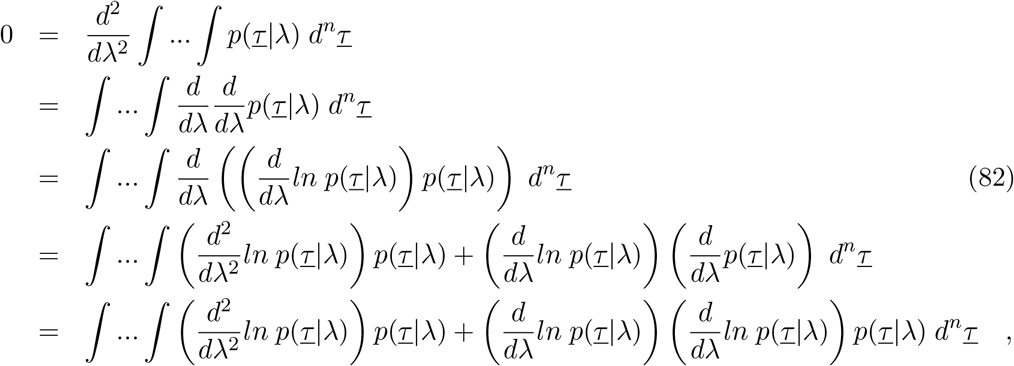

and hence

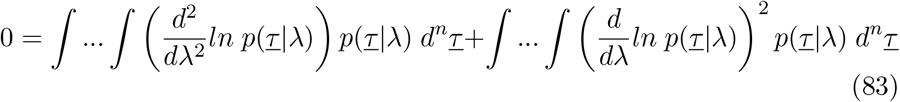

Or

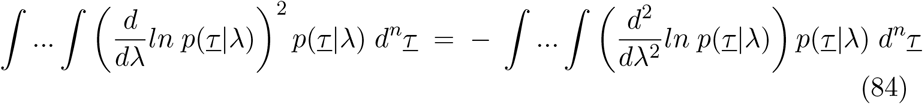

with the Cramér-Rao inequality now in its final form given by

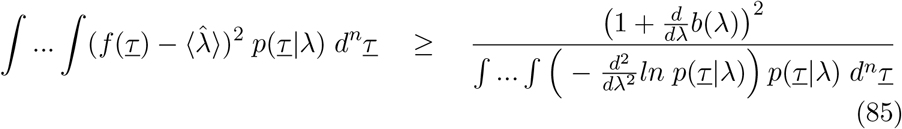

with the Fisher information

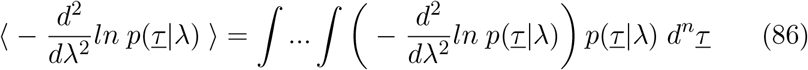

in its expectation value form in the denominator of the inequality and the variance

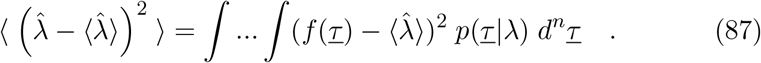

So we can write in short

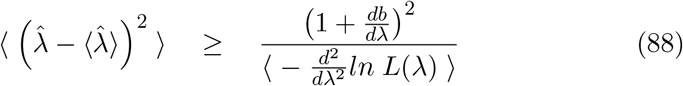

as the easiest memorizable form of the Cramer-Rao inequality, using the likelihood notation *L*(*λ*) = *p*(<u>*τ*</u> |*λ*), since the ensemble average is always taken over the data points <u>*τ*</u> now.

#### B.2 Application to the exponential distribution and its maximum likelihood estimator

We first calculate explicitly the ensemble mean of the estimator by deriving an ODE and solving it. From Eq. (70) we have with 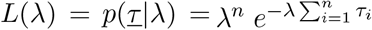 and 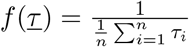 for the exponential distribution

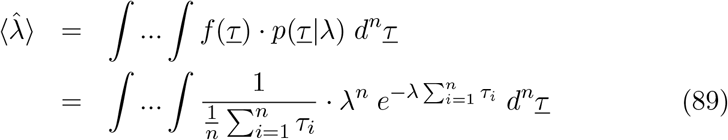

and hence, for its derivative

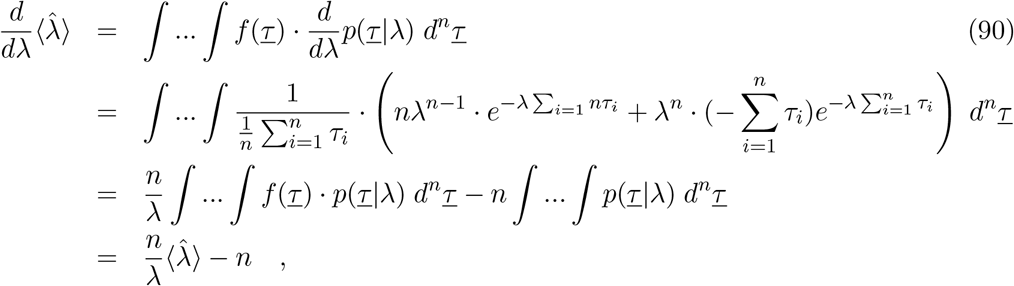

since the first integral 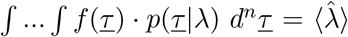 gives the expectation value, and the second ∫ … ∫*p*(<u>*τ*</u> |*λ*) *d*^*n*^<u>*τ*</u> = 1 gives the normalization of probability. Thus, we have the ODE for 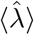 given by

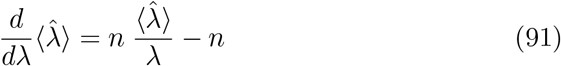

to be solved. This can be done via the variable transformation 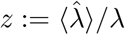, hence 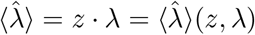. We transform the original ODE into an ODE in new variable

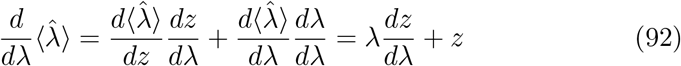

and on the other side of the original ODE

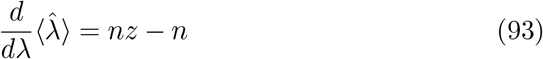

to be solved

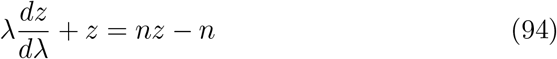

Or

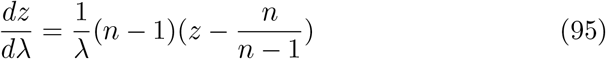

which can now be resolved easily via separation of variables. Then,

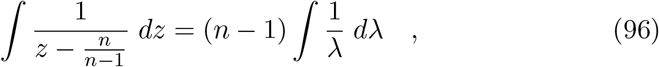

giving as general solution

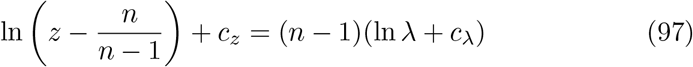

and after some elementary steps, and replacing *z* with 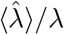 again, we have

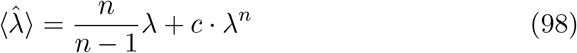

using integration constants *c*_*z*_ and *c*_*λ*_ to be combined to 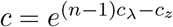. To leading order in *λ* we have the solution 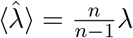, since *λ* is in general a small number.

Hence, for the bias 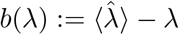 we have

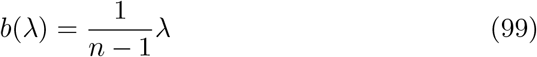

and in the expression for the Cramér-Rao bound of the ensemble variance

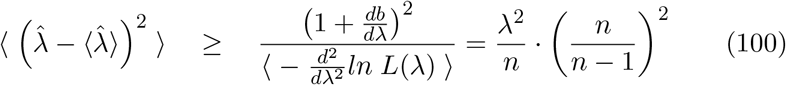

since the expectation of the negative second derivative of the log-likelhood function is 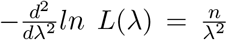 independent of the data *τ*, as calculated previously. Therefore, the expectation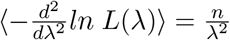, also know as the Fisher information).

In conclusion, the analysis of the maximum and its vicinity of the likeli-hood using the empirical data gives the same information as the ensemble analysis of mean and variance of the estimator, just with small extra terms which vanish for large data sets. Already for 10 data points, (for instance using the Gaussian approximation), compared with 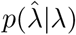 it can be quite good, and gives qualitatively the correct result for confidence interval analysis (see for another example [47] in Figure 2 b)), using extended simulations for the conditioned probability of estimators from the original parameter.

